# Integrated Single Cell and Spatial Transcriptomics Reveal Autoreactive Differentiated B Cells in Joints of Early Rheumatoid Arthritis

**DOI:** 10.1101/2021.08.09.21260015

**Authors:** Uta Hardt, Konstantin Carlberg, Erik af Klint, Peter Sahlström, Ludvig Larsson, Annika van Vollenhoven, Susana Hernandez Machado, Lena Israelsson, Khaled Amara, Karine Chemin, Marina Korotkova, Gunilla B Karlsson Hedestam, Anca I Catrina, Sarah A Teichmann, Patrik L Ståhl, Vivianne Malmström

## Abstract

Rheumatoid Arthritis (RA) is a prevalent autoimmune disease characterized by inflammation of peripheral joints. Patients can be subdivided by the presence or absence of Rheumatoid Factor and anti-citrullinated protein antibodies (ACPA) in their circulation. Inflammation of the joint tissue is associated with infiltration of leukocytes from the blood, which can result in generation of lymphoid structures composed of B and T cells. Previous studies have shown that both memory B cells and antibody-secreting plasma cells populate the rheumatic joint tissue when captured from established and often long-standing disease. However, it has remained unclear, whether these cells are autoreactive and whether the associated lymphoid structures are present at the site of inflammation already at the time of diagnosis. Here, we used an integrated single cell and spatial transcriptomic approach to study B and plasma cells in synovial tissue of ACPA- and ACPA+ RA patients at this early time point. We found evidence for T cell help to B cells and presence of memory B and plasma cell pools in ACPA- as well as in ACPA+ RA. Our results demonstrated common supportive microenvironments in both patient subgroups, clonal relationships between the memory B and plasma cell pools and autoreactivity within the plasma cell compartment. These findings challenge our understanding of the dynamics of local adaptive immune responses in the RA joint of ACPA- and ACPA+ patients at the time of diagnosis, with direct implications for B and T cell targeting therapies for both patient subgroups.

**One Sentence Summary:** A B cell maturation and plasma cell maintenance niche is evident at onset of Rheumatoid arthritis in both ACPA- and ACPA+ patients.

## INTRODUCTION

Rheumatoid arthritis (RA) is a chronic autoimmune disease characterized by inflammation of peripheral joints *(1)*. Two thirds of the patients can be further distinguished by occurrence of autoantibodies in circulation *(2)*. These autoantibodies include Rheumatoid Factor, *i*.*e*. an anti-Fc gamma of IgM or IgA isotype, and anti-citrullinated protein antibodies (ACPA) that are clinically assessed by an anti-cyclic citrullinated peptide (anti-CCP) IgG ELISA. The autoantibodies often co-exist and can precede the clinical onset of RA by years *(3–5)*. The synovial tissue of RA patients displays varying degrees of inflammatory infiltrates, which can be broadly subcategorized into three histological pathotypes: scarcely infiltrated by immune cells, diffusely infiltrated by immune cells or harboring ectopic lymphoid structures *(6)*. These lymphoid structures comprise T and B cells which can contribute to HLA class II-dependent immune responses that are associated with ACPA+ disease *(7)*. While such immunological features in the inflamed synovia are well recognized in long-standing ACPA+ RA, it is unknown whether *in situ* B cell differentiation into autoantibody producing plasma cells occurs already at the onset of RA. Here, we studied joint biopsies from ACPA+ and ACPA-RA patients using a cutting edge integrated single cell and spatial transcriptomic approach, coupled with bioinformatics analysis, to determine whether the affected tissue harbored B cells with plasma cell differentiation potential at the time of diagnosis. We further analyzed the properties of memory and antibody-secreting plasma cells in their local niche. Finally, we investigated the clonality of synovial plasma cell-derived monoclonal antibodies within the memory B cell compartment, and examined their reactivity towards disease-associated antigens.

## RESULTS

### Subhead 1: Phenotype and tissue context of synovial B cells at the onset of RA

Eight synovial tissue biopsies from small (MCP, wrists) and large (knee) joints from four ACPA+ and four ACPA-RA patients collected within two days of diagnosis were included in the study. A summary of the clinical features of the patients is displayed in Supplemental Table 1. Following flow cytometric sorting for CD3-CD14-AnnexinV-CD19+ cells, we retrieved viable B cells from all samples (Figure 1A). The amount of tissue was critical, as very few B cells (16 and 24) were recovered from 2 of the 3 small joint samples, while the arthroscopy material from knee joints yielded numerous B cells (400-800) in 4 of the 5 samples (Figure 1B). We subdivided the isolated B cells based on IgD and CD27 protein expression. Hereby, 20-45% of the B cells displayed a naïve IgD+CD27-phenotype, while 24-63% were of a IgD-CD27+ classical memory phenotype, 1-4% were CD27++, 2-4% had an IgD+CD27+ unswitched memory phenotype and 8-29% had an IgD-CD27-phenotype (Figure 1C). These subsets were apparent irrespective of the ACPA status of the patients.

**Fig 1.**
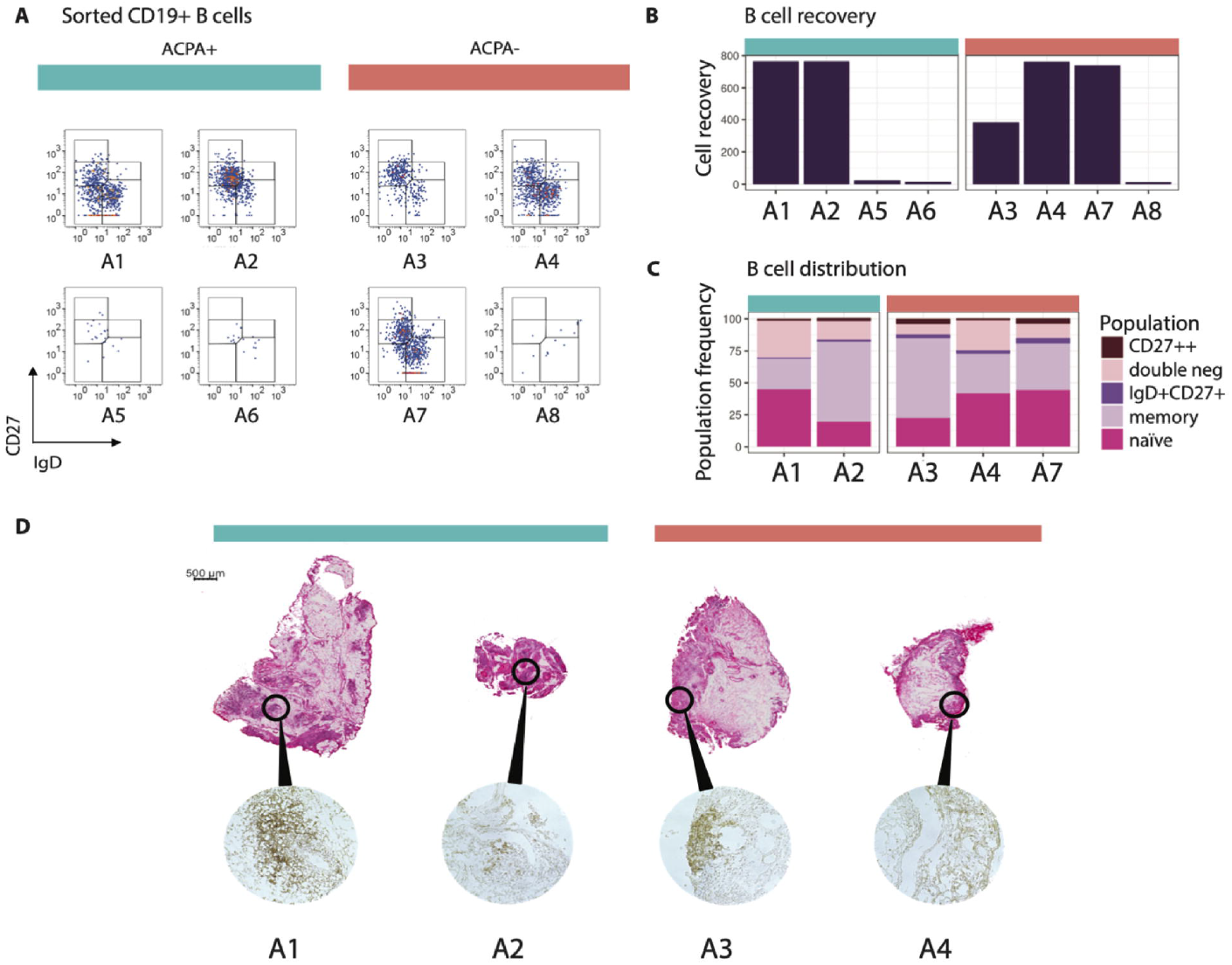
Number, phenotype and spatial distribution of synovial B cells in Rheumatoid Arthritis. **(A)** The flow cytometry panels show all CD3-CD14-AnnexinV-CD19+ single B cells from synovial biopsies of 4 ACPA+ and 4 ACPA-RA patients at the time of diagnosis in pseudocolor plots, where red color indicates data point overlap. A5 was taken from a metacarpophalangeal (MCP) 4 joint, A6 and A7 were taken from wrists. Remaining synovial biopsies are arthroscopic biopsies from knees. **(B)** 5 out of 8 biopsies exceeded a minimum of 382 B cells suitable for phenotypic characterization and scRNAseq analysis. **(C)** Memory B cells made up the largest population of synovial B cells (24-63%), while naïve cells were a bit fewer (20-45%). Double negative B cells had a variable range (8-29%), while a minority was CD27++ (1-4%) or of the unswitched IgD+CD27+ memory phenotype (2-4%). **(D)** H&E-staining shows the tissue architecture of A1-A4. Immunohistochemistry staining with anti-CD19 and anti-CD20 antibody cocktail confirms the presence of B cells.

Four biopsy samples (A1-A4) were available for further studies focusing on the inflammatory architecture. H&E-staining and immunohistochemistry (Figure 1D) showed scattered B cell-rich infiltrates, especially surrounding vessels, in samples A1 and A3, while it showed more dispersed B cells in samples A2 and A4. The analysis validated the presence of B cells in all studied biopsies with no obvious difference in B cell density between the two patient groups.

### Subhead 2: Single cell and spatial transcriptome analyses of synovial B cells

Altogether, we index-sorted 3,468 individual B cells (14-764 cells/donor) from synovial biopsies from eight RA patients using flow cytometry (Supplementary Figure 1). Five out of the eight samples had sufficient B cell numbers (>382 cells) and were further processed for scRNAseq. Three out of the five passed initial QC; these originated from one ACPA+ and two ACPA-RA patients. In uniform manifold approximation and projection (UMAP) embedding, the B cells from patients A2-A4 distributed into three clusters. We identified 113 plasma cells (6%), 395 naïve B cells (21%) and 1388 memory B cells (73%, Figure 2A, Supplementary Figure 2).

**Fig 2.**
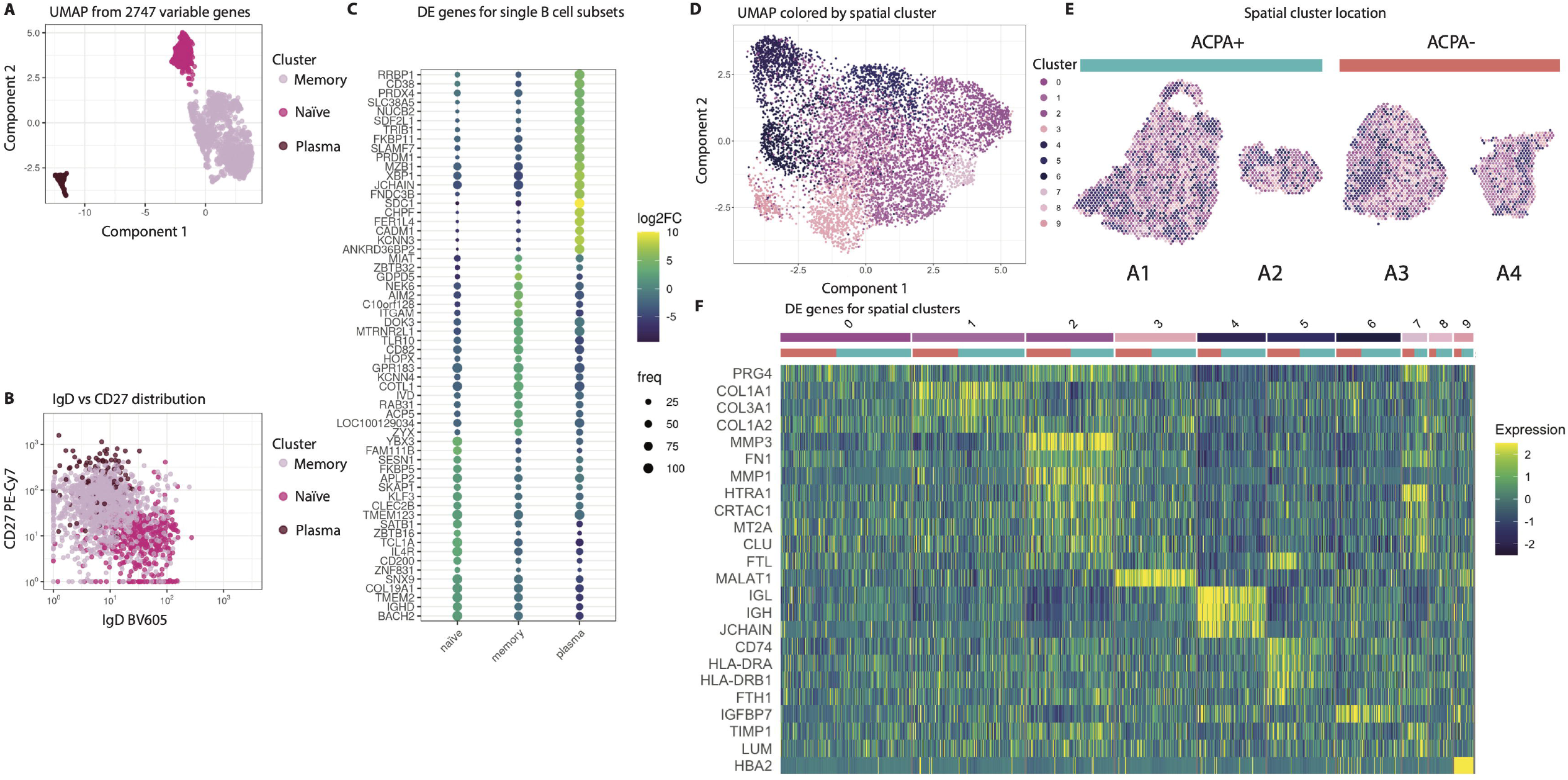
Identification of synovial B cell clusters and assessment of context relevant markers. **(A)** The UMAP dimensionality reduced B cell transcriptome from A2-A4 reveals the presence of a naïve, a memory and a plasma cell cluster. **(B)** The scRNAseq annotation translates back onto the expected phenotypic marker expression by flow cytometry. **(C)** Average log2FC and robustness of expression for differentially expressed genes of B cell transcriptomes show marker genes for naïve, memory and plasma cells. **(D)** UMAP on spatial transcriptomics data colored with the ten nearest neighbor clusters. **(E)** Clusters are shown on the tissue sections. **(F)** Differentially expressed genes between the ten clusters. Cluster 4 shows genes strongly associated to plasma cells.

Assignment of the cluster labels from the scRNAseq back into the flow cytometry distribution of IgD vs CD27 recapitulated the index labels from the sort validating our approach (Figure 2B, Supplementary Figure 3-4). Figure 2C depicts the top-20 most differentially expressed genes (DEGs) between the three clusters. The most distinct features expressed by the plasma cell population included the plasma cell markers *XBP1, SDC1* (encoding CD138), *SLAMF7* and *PRDM1* (encoding BLIMP-1). Additionally, this cluster had increased expression of *JCHAIN*, an indication of high antibody protein production. In contrast, the memory B cell cluster was characterized by *ITGAM* encoding CD11b and *GPR138* encoding EBI2, which is typical of memory B cells. The naïve B cell cluster was characterized by *IGHD* and *IL4R* expression encoding IgD and IL4 receptor which are representative of naïve resting B cells.

Two ACPA- and two ACPA+ tissues were processed also for spatial transcriptomics. Hereby, we generated mRNA data on 18,199 genes over 7334 spots distributed on several consecutive cryosections. The number of transcripts/spot was heterogeneous as expected from the tissue morphology observed in the H&E-staining. We verified that RNA quality was adequate in less cell dense areas by assessing spatial RIN scores (Supplementary Figure 5). After data integration and subjection to UMAP, we identified 10 clusters (Figure 2D) that spatially mapped into the sections (Figure 2E). These clusters matched the different morphological areas (Figure 1D). The dark blue clusters (4, 5 and 6) overlapped with the cell dense lymphocytic areas and were present in all four patients. The heatmap (Figure 2F) shows the top DEGs of the 10 clusters, to which both the ACPA+ and ACPA-samples contributed. We additionally subjected the data to a PCA visualizing that the patient subgroups were largely overlapping (Supplementary Figure 6).

Cluster 4, which was in proximity of the infiltrate regions and the B cell-rich areas had a strong expression of *JCHAIN*, IGH, IGL and *MZB1, SDC1* and *XBP1*, suggesting the presence of plasma cells in these locations (Supplemental Table 2). Cluster 5 had a strong signal for immune cells with high expression of HLA class II genes and *CD74* indicating antigen presentation.

Close by cluster 4 and 5 was cluster 6 with a weaker immune signature although containing genes encoding the chemokine CCL19 known to attract B and T cells.

The remaining clusters overlapped with areas outside the infiltrates, and contained genes associated to connective tissue such as collagen, metallopeptidases, fibronectin and proteoglycan as well as some immunological features such as complement factor C3. This analysis revealed the heterogeneity of the tissue even in seemingly homogenous morphological areas (Supplemental Table 2).

### Subhead 3: B-T cell interaction in the RA joint tissue

All B cell subsets expressed high amounts of *CD74* encoding the HLA invariant chain as well as the *HLA-DRB1* and the paired *HLA-DRA* chain genes implicating antigen-presentation capacity (Figure 3A, Supplementary Figure 7). Based on this observation, we further investigated other B cell transcript expressions indicating T cell interaction. We found expression of the genes encoding the costimulatory molecules CD40, CD80, CD86, CD84 and also low expression of *TNFSF4* encoding CD134 (also known as Ox40L). Furthermore, primarily naïve B cells expressed *ICOSLG, IL4R* and *IL21R*. IL21 receptor signaling is important for B cell proliferation and class switch recombination as well as for plasma cell differentiation. Robust expression was also found for *IL16*, which encodes a cytokine ascribed to be a CD4 T cell chemoattractant.

**Fig 3.**
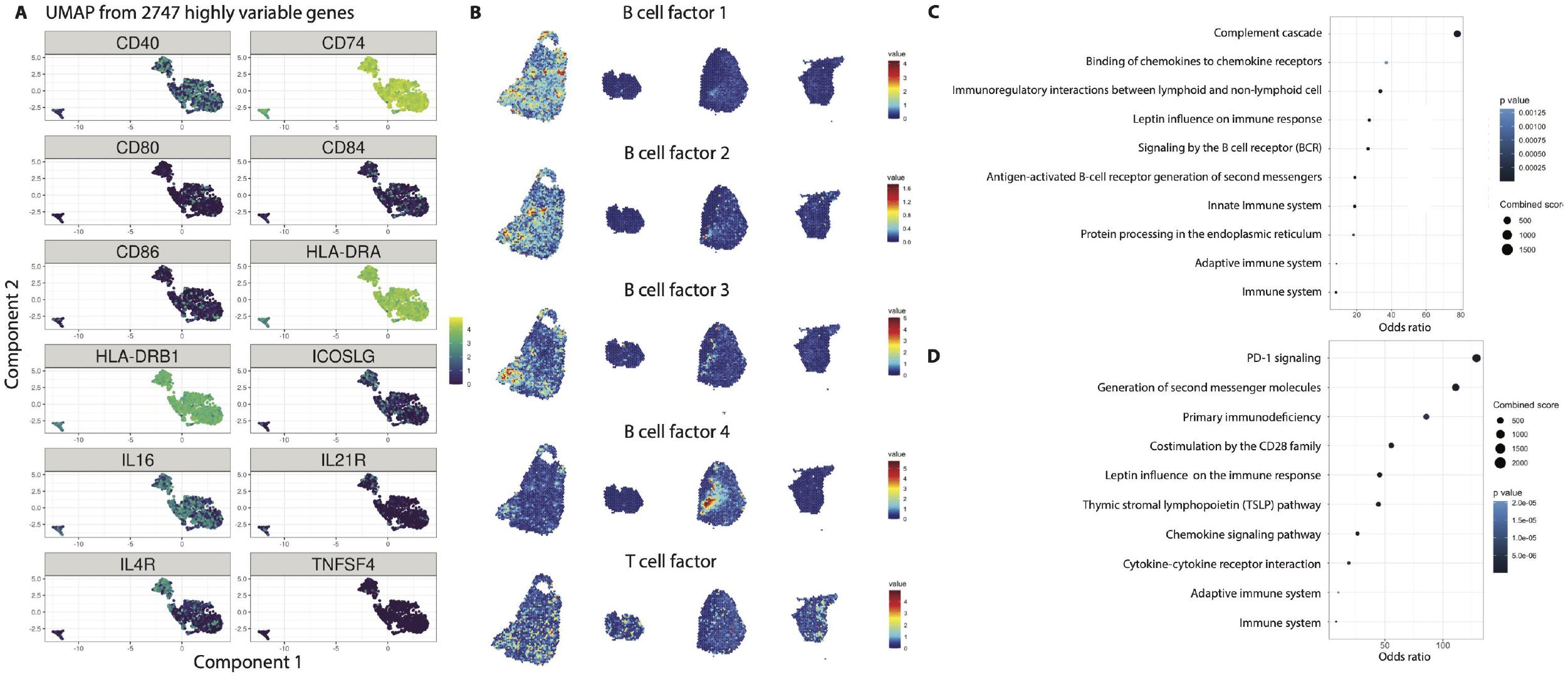
T cell interaction as a means of B cell maturation in the synovial tissue. **(A)** We found increased expression of *HLA-DRB1, HLA-DRA* and *CD74* antigen presentation genes as well as expression of costimulatory molecules that are involved in B-T cell interaction, such as *CD40, CD80* and *CD86*, as well as *IL21R*, which favors plasma cell differentiation. **(B)** Four B cell factors and one T-B cell interaction factor are shown with gene activities overlapping different morphological areas. **(C)** Pathway analysis from the top-20 genes for B cell and **(D**) T-B cell interaction factors (T cell factor) in (B).

From the tissue transcriptional data, factor analysis was performed, and 20 different factors and their driver genes could be identified (Supplementary Figure 8). Five factors showing a B cell and T-B cell interaction signature (6,5,8,13,17) are presented in Figure 3B. The associated genes underwent pathway analysis through NCATS BioPlanet 2019 (Figure 3C-D). This analysis revealed BCR signaling for the factors denoting increased B cell activity and PD-1 signaling as the top hit in the T-B cell interaction pathways. Again, both the ACPA+ and ACPA-biopsies displayed clear signals of B-T cell crosstalk.

### Subhead 4: Transcriptomic profile and spatial context of memory B cells

All memory B cell-associated markers were either shared with naïve B cells or with plasma cells (Figure 4A, Supplementary Figure 9). *CCR6, CR2* (encoding CD21), *FCER2* (encoding CD23), *ITGB2* (encoding CD18) and elevated expression of *MS4A1* (encoding CD20) were shared with the naïve B cell cluster, while *TNFRSF13B* (encoding TACI), *CD27* and *CXCR3* expression was shared with the plasma cell cluster. Also, gene expression of the transcription factors IRF8, PAX5 and Spi-B as well as the transcription co-regulator SKI were shared with the naïve B cell cluster.

**Fig 4.**
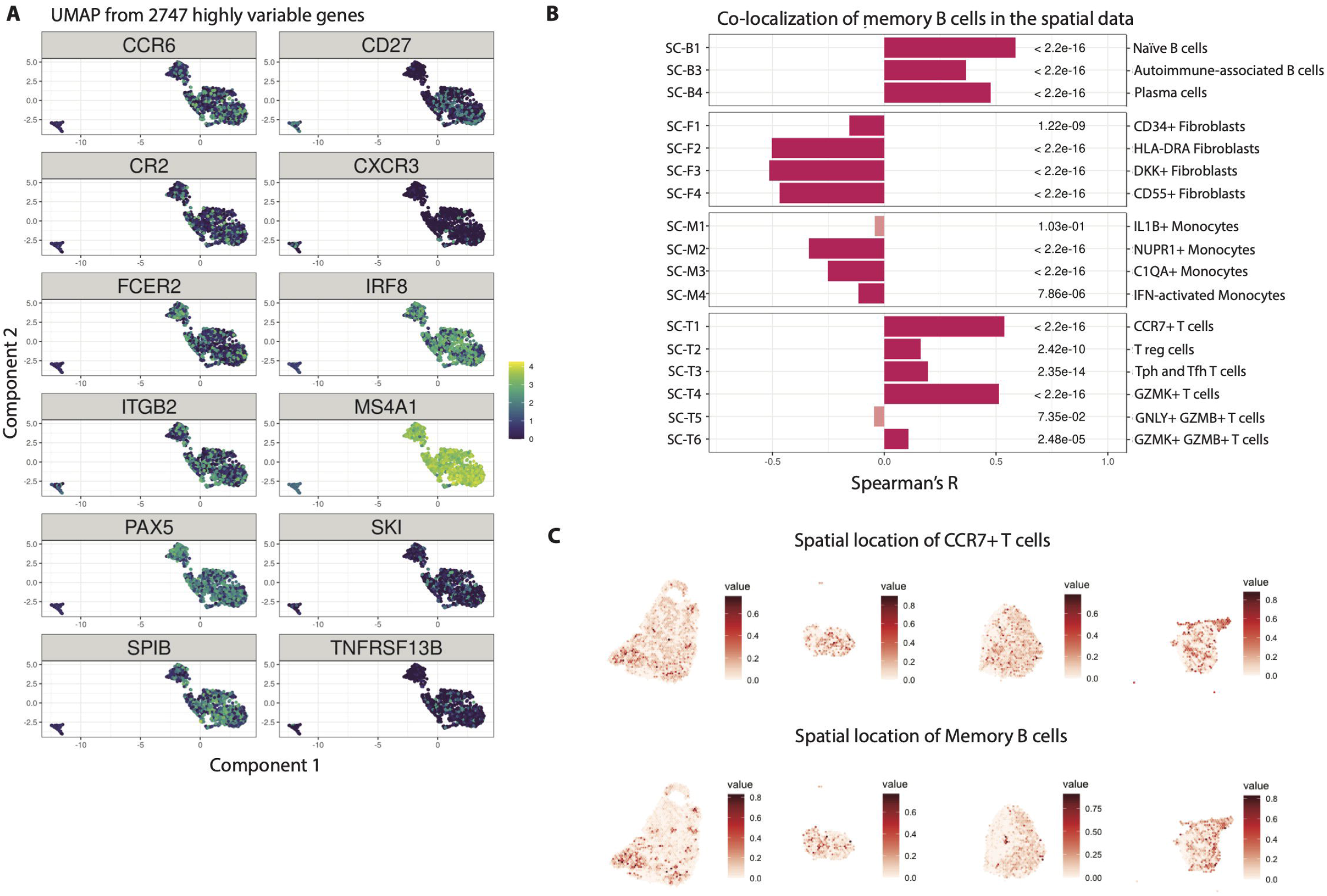
Memory B cell compartment associated with lymphocytic infiltrates in synovial tissue. **(A)** All the memory cell markers are either shared with the naïve cell cluster, such as *CCR6, CR2* encoding CD21, *FCER2* encoding CD23 or the transcription factors *IRF8* and *PAX5*, or they are shared with the plasma cell cluster, such as *CD27* and TACI encoded by *TNFRSF13B*. **(B)** We determined correlation between the predicted score for the location of the memory B cell (SC-B2) cluster and the predicted scores for the location of the other single cell clusters containing other B cell subsets, fibroblasts, monocytes and T cells in the spatial data *(8)*. **(C)** The spatial distribution of memory B cells (SC-B2) and *CCR7*+ T cells (SC-T1) on the sections show a dispersed but correlating pattern.

Next, we investigated the spatial context of memory B cells. In order to enrich our findings, we used previously published single cell signatures by Zhang et al. *(8)*. Similarity of our and their memory B cell (SC-B2) signature was validated by Fisher’s exact test (OR=99.6, p-value<2.2e-16, Supplemental Table 3). We then used Spearman’s R to assess co-localization of memory B cells (SC-B2) with the other described single cell subtypes (Figure 4B-C, Supplementary Figure 10). Memory B cells (SC-B2) mostly co-occurred with other B cells (0.37≤ R≤ 0.59) suggesting clonal expansions, and T cells (0.11≤ R≤ 0.54, Figure 4B) suggesting T cell interaction. Notably, the memory B cells (SC-B2) co-localized with the T peripheral or follicular helper cell subset (SC-T3, R=0.20), but also with *CCR7*+ T cells (SC-T1, R=0.54) and *GZMK*+ T cells (SC-T4, R=0.51). There was an inverse correlation for co-localization with most fibroblast (−0.51≤ R≤ -0.16) and monocyte (−0.34≤ R≤ -0.12) subsets.

### Subhead 5: Transcriptomic profile and spatial context of plasma cells

We found terminally differentiated plasma cells in the rheumatoid synovium of both ACPA+ and ACPA-RA patients at the time of diagnosis. Besides the top-20 DEGs (Figure 2C), plasma cells were further distinguished by expression of genes that are important for matrix interaction, such as *ICAM2* and *ITGA6* encoding CD49f (Figure 5A, Supplementary Figure 11). Additionally, plasma cells expressed *CCR2, CCR10* and *CXCR4*, which is important for the CXCL12-dependent plasma cell survival niche. Still, *CXCR4* expression was similarly to *IRF4* expression not exclusive to plasma cells. A gene that was more exclusive to plasma cells and also mediates plasma cell survival is *TNFRSF17* encoding the APRIL receptor BCMA. Additional markers that associated with plasma cells include *IL6R, PECAM1* (encoding CD31), *SELPLG* (encoding CD162) and the large neutral amino acid transporter, CD98, formed by the gene products of *SLC3A2* and *SLC7A5*.

Similar to the analysis for Figure 4, we took advantage of the previously published scRNAseq signatures *(8)* to describe the location and context of plasma cells in our spatial transcriptomic data. The Fisher’s exact test validated the overlap between the DEGs in our plasma cell cluster and the previously described plasma cell (SC-B4) cluster (OR=45.1, p-value<2.2e-16, Supplemental Table 4). As memory B cells (SC-B2) and plasma cells (SC-B4) were highly co-localized (R=0.48), we observed a similar pattern for co-localization for plasma cells (SC-B4) as for memory B cells (SC-B2, Figure 5B). Plasma cells (SC-B4) co-localized with other B cell subsets (0.23≤ R≤ 0.48) and several subsets of T cells namely CCR7+ T cells (SC-T1, R=0.42), T peripheral or follicular helper cells (SC-T3, R=0.13) and GZMK+ T cells (SC-T4, R=0.33). In contrast, the Spearman correlation was inverse for GNLY+ GZMB+ T cells (SC-T5, R=-0.07), fibroblasts (−0.47≤ R≤ -0.17) and monocytes (−0.34≤ R≤ -0.11). Spatially, the plasma cell (SC-B4) cluster was more distinct than the memory B cell (SC-B2) cluster (Figure 4C, Figure 5C, Supplementary Figure 12). The likelihood of finding plasma cells in the tissue was increased in the vicinity of lymphocyte infiltrates as also the correlation estimates were somewhat lower between plasma cell (SC-B4) and B or T cells compared to memory B cell (SC-B2) and B or T cells. Moreover, we observed a high overlap with the predicted location for plasma cells (SC-B4) and the spatial cluster 4 (Figure 2F), which we concluded to have a plasma cell signature (Figure 5D). Additionally, we found a high overlap with *CXCL12* expression in the tissue (Figure 5E).

**Fig 5.**
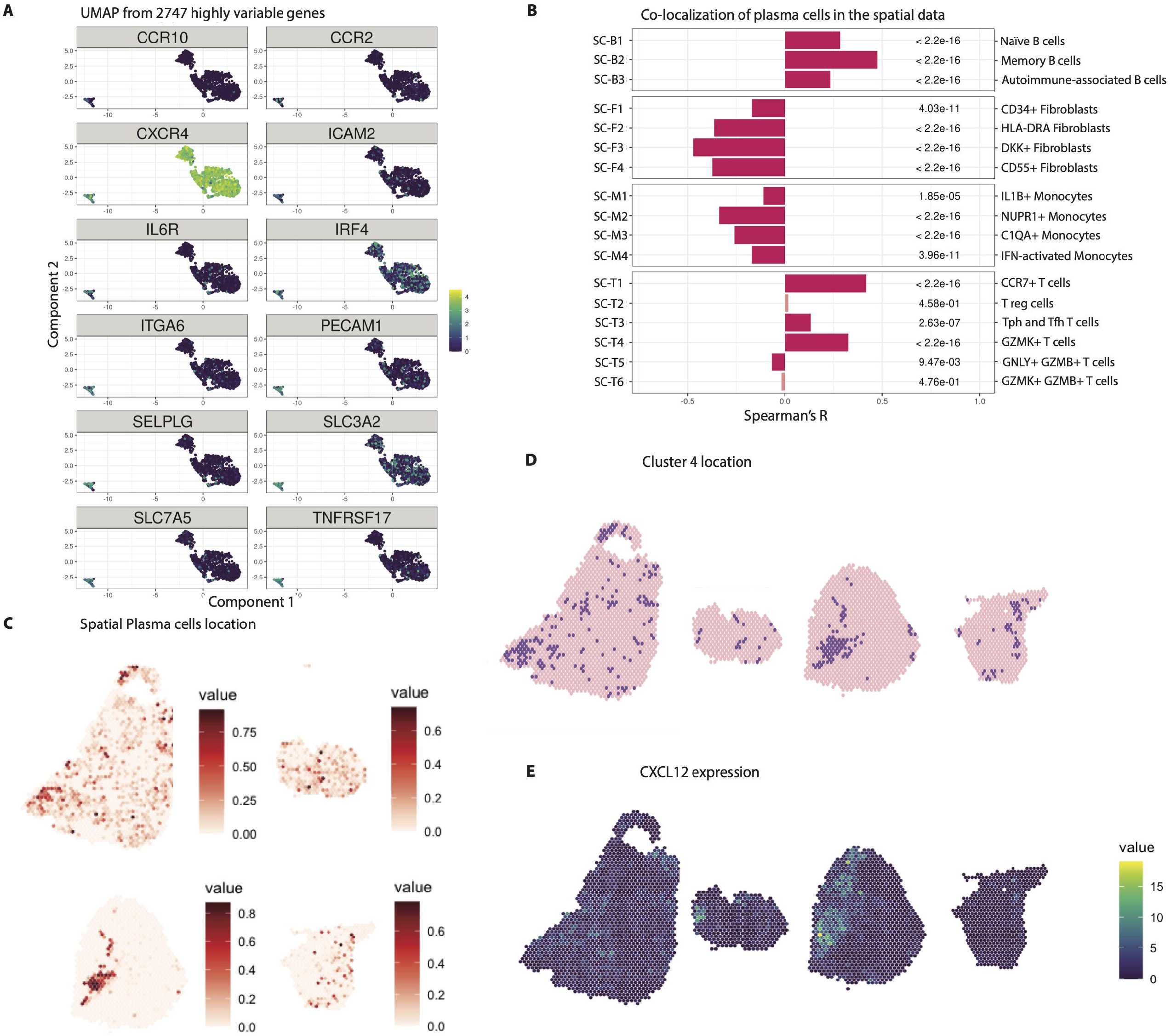
Plasma cell compartment and niche in the synovial tissue at the time of RA diagnosis. **(A)** Besides the classical markers identified in the differential gene expression analysis, we find expression of genes in plasma cells that are important for matrix interaction such as *ICAM2* and *ITGA6* encoding CD49f. We also find expression of the chemokine receptors *CCR2, CCR10* and vast expression of *CXCR4*, which is important for plasma cell maintenance. Still, *CXCR4* expression was similar to *IRF4* expression not exclusive to plasma cells. Moreover, we detect the expression of BCMA encoded by *TNFRSF17*, which is also important for the plasma cell niches. **(B)** We determined correlation between the predicted score for the location of the plasma cell (SC-B4) cluster and the predicted scores for the location of the other single cell clusters containing other B cell subsets, fibroblasts, monocytes and T cells in the spatial data *(8)*. **(C)** Spatially, plasma cells seem to be confined and adjacent to the lymphocyte infiltrates. **(D)** We find overlap between the designated plasma cell cluster 4 and the predicted plasma cell location by the single cell data. **(E)** Additionally, we find high CXCL12 expression in the plasma cell areas suggesting the existence of a plasma cell niche in the inflamed tissue.

The CXCL12-CXCR4 axis is indispensable for plasma cell maintenance and our results suggest that this axis is active already at the time of diagnosis.

### Subhead 6: Expression, clonality and reactivity of plasma cells

Of the 113 identified plasma cells and their scRNAseq data, we reconstructed the full BCR sequence and expressed antibodies corresponding to 9 unique clones, 3 clones with 2 cells each, 1 clone with 4 cells, 1 clone with 5 cells and 1 clone with 21 cells (Figure 6A, Figure 6B, Supplementary Figure 13). Additionally, 9 of these 15 clones were found to have sibling cells with an identical variable and constant chain pair within the memory B cell compartment. The identified clonotypes of biopsy A3 appeared to be less polyclonal than those of A2 and A4. For the largest clone, we ruled out potential index-hopping that might mis-assign BCR sequences to neighboring wells (Supplementary Figure 14). The 5 clonotypes of biopsy A2 comprised 70 out of 753 memory and plasma cell members, the 2 clonotypes in A3 comprised 82 and 32 of 371 memory and plasma cell members and the 7 clonotypes of biopsy A4 comprised 38 out of 377 memory and plasma cell members, respectively. The expressed antibodies hence originated from all three biopsies with 5 antibodies derived from A2, 2 from A3, and 8 from A4.

**Fig 6.**
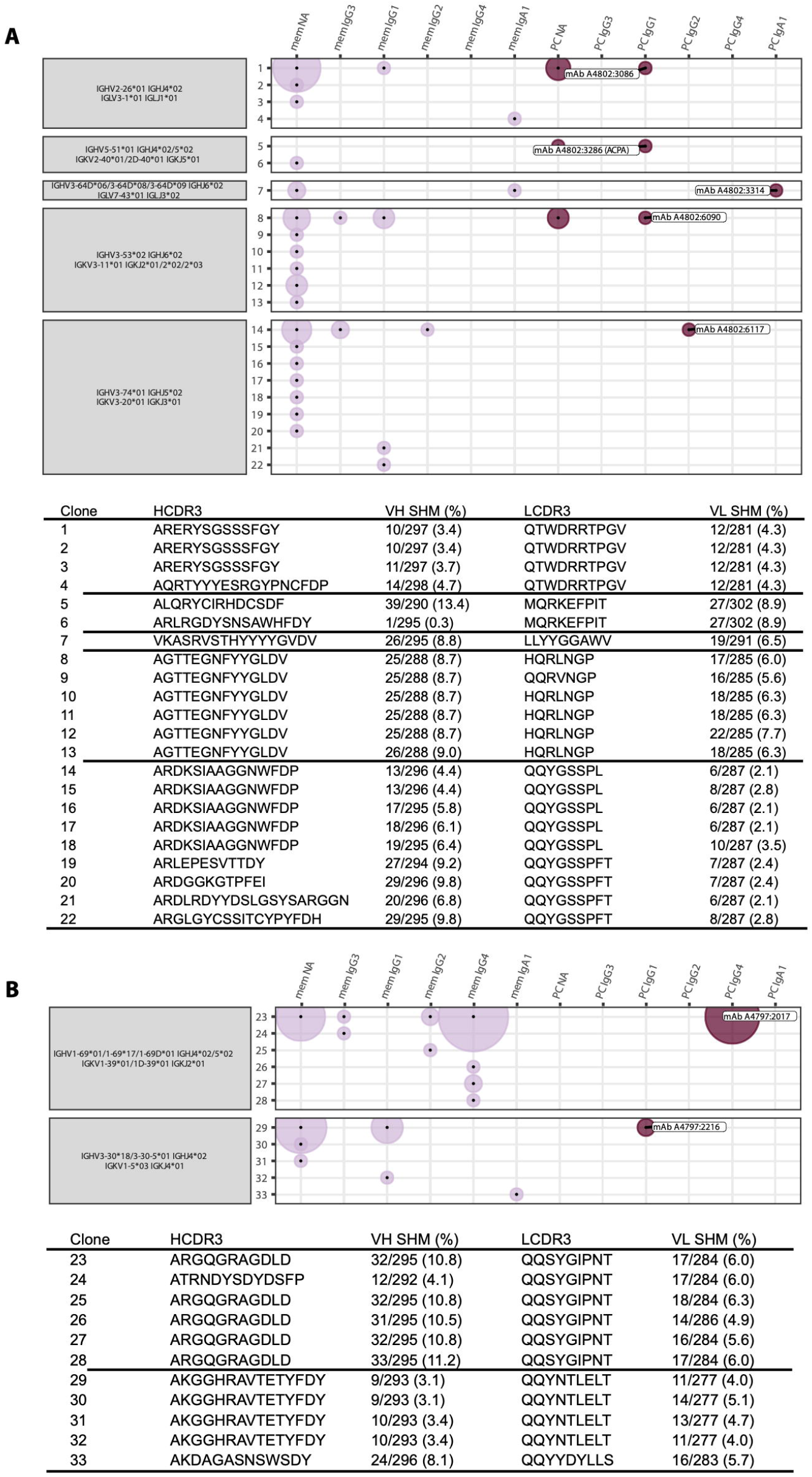
Clonotype characterization of synovial plasma cell derived antibodies. **(A)** The plot shows clonotype information for five plasma cell derived monoclonal antibodies from A2. Each clonotype is defined by identical heavy and light chain V and J gene usage (grey boxes). Each clone with a unique sequence for the combination of heavy and light chain is assigned a number on the y-axis. This sequence is further described by the number of SHM and the complementary determining region 3 (CDR3) for the heavy and light chain in the table below the plot. The x-axis and dot color depict the memory B (mem) or plasma cell (PC) cluster identity of the clone members by transcriptomic profile. The x-axis additionally defines the heavy chain isotype of the identified clone members in the order of possible class switch recombination from left to right. NA means not available from the data. The area of the dots is proportional to the number of identified members; e.g. the largest area within the first clone is equivalent to 16 members while the smallest area is equivalent to 1 member. **(B)** The plot shows the equivalent clonotype information that is shown in panel (A) for the two plasma cell derived monoclonal antibodies originating from A3.

Figure 7A shows the 5 antibody clonotypes from the ACPA+ individual embedded in a phylogenetic tree with all recovered paired full length BCR sequences from that individual. All 5 clonotypes originated from expanded clones. Two of the recombinantly expressed monoclonal antibodies from the ACPA+ patient, A4802:3286 and A4802:3314, gave positive signals in the anti-CCP2 ELISA, while the other antibodies were nonreactive (Figure 7B). However, A4802:3314 (an IgA1 and IgL3 clone) was further positive in the poly-reactivity ELISA and was therefore not defined as an ACPA. To continue investigate the reactivity of the A4802:3286 ACPA clone, we examined its potential multi-reactivity towards other post-translational modifications using the Orgentec clinical test containing a mutated vimentin backbone peptide. Here, the citrulline reactivity was positive down to a concentration of 2ng/ml (Figure 7C, Supplementary Figure 15). In contrast, this clone was negative for other commonly studied citrulline specificities on a multi-peptide array (Supplementary Figure 16). We examined the level of gene expression and found abundant expression of vimentin on the patient specific section (Figure 7D).

**Fig 7.**
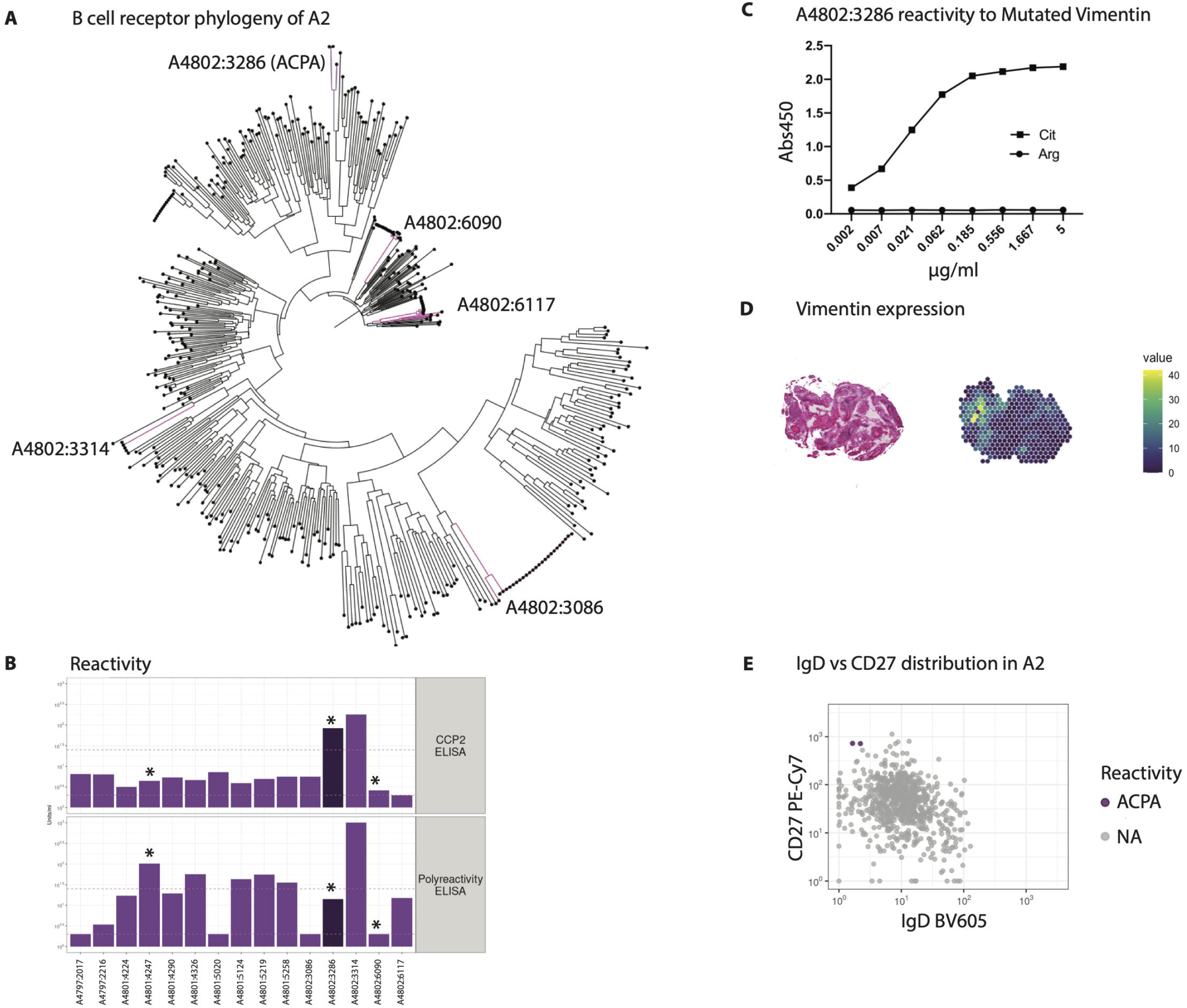
Reactivity landscape of synovial plasma cell derived antibodies. **(A)** Phylogenic tree of the B cell receptor clones of one of the ACPA positive individuals (A2). Clonotypes from figure 6 are highlighted. **(B)** One of the monoclonal antibodies was identified as an anti-citrullinated protein antibody (ACPA) based on being CCP positive, but not polyreactive (dark purple). Several antibodies showed polyreactivity. * indicates insertion of N-linked glycosylation sites by SHM. **(C)** The A4802:3286 ACPA has strong reactivity towards mutated citrullinated vimentin peptide. **(D)** Vimentin is expressed within the tissue slides analyzed by spatial transcriptomics. **(E)** The A4802:3286 ACPA derives from two IgD-CD27++ plasma cells.

The A4802:3286 ACPA clone was of the IgK and IgG1 isotypes and originated from two CD27++IgD-plasma cells in the original flow cytometry data (Figure 7E). Furthermore, somatic hypermutation (SHM) had introduced two glycosylation sites in the antigen binding fragment (Figure 7B, Supplementary Figure 17), which is a common feature amongst ACPAs. Out of the studied plasma cell clones, the A4802:3286 ACPA also had the highest level of SHM for both the heavy and light chains with 39 and 27 mutations, respectively (Figure 6A). The ACPA clonotype used IGHV5-51*01, IGHJ4*02/5*02, IGKV2-40*01/2D-40*01 and IGKJ5*01 and had 3 members in total; one identical plasma cell sibling and one similar memory cell member with an identical light chain and a similar heavy chain. To summarize, we describe a monoclonal ACPA derived from two identical plasma cells from synovial tissue of early RA with a narrow reactivity pattern to a citrullinated RA antigen.

## DISCUSSION

Here, we investigated the B cell compartment of joint biopsies from two ACPA+ and two ACPA-RA patients obtained within two days of diagnosis. We visualized the localization of B cell subsets within the biopsies and characterized them with regard to both T cell crosstalk and plasma cell survival niches. Moreover, we investigated B cell clonotypes shared between the memory and plasma cell subsets and we identified ACPA reactivity in the plasma cell compartment.

In our unique approach, we integrated single cell resolution and spatial transcriptomic data from patients at the time of diagnosis, while previous studies *(8–10)* analyzed single cells from dispersed synovial tissue from RA or osteoarthritis patients at mixed time points of the disease. We embedded parts of these studies in our analysis to enrich our findings with respect to the memory B and plasma cell context. In line with the study by Zhang et al. *(8)* and Scheel et al. *(9)*, we also found that not all patients presented with high amounts of leukocytes or specifically B cells in the biopsy. In our experience, B cell yields correlated with disease activity at the time of sampling as well as with amount of tissue, but was independent of ACPA status.

We describe the presence of naïve, memory and plasma cells in the disease-affected synovial tissue of both ACPA+ and ACPA-RA patients. This is intriguing since it has been hypothesized that ACPA+ RA patients would have a stronger B-T cell drive in their disease based on the genetic HLA-DR association and the presence of autoantibodies. Our data instead demonstrate that the inflamed joint is an attractive site for B cells and plasma cell differentiation, also in ACPA-RA patients. Indeed, while ACPA+ RA patients respond better to B and T cell targeted therapies, ACPA-RA patients can also benefit from such therapeutic interventions *(11, 12)*.

Based on our transcriptomic data, we could identify many shared features between ACPA+ and ACPA-RA patients at this early time point before therapeutic intervention. Importantly, we also found citrulline autoimmunity within the ACPA+ RA joint and we elaborate on a local immunological B cell maturation process via T cell help as previously suggested by Rao et al. *(13)*. Additionally, we describe the nature of memory B cells and plasma cells in the joint and find supporting niche structures. Our data suggest BAFF and APRIL dependent survival of terminally differentiated B cells at the site of inflammation already at the time of RA diagnosis. Additionally, we find strong CXCR4 expression in memory B and plasma cells and elevated CXCL12 expression in the supporting tissue. This supporting niche has not been previously described to be maintained at the site of joint inflammation, but is well known to be crucial for plasma cell survival in the bone marrow *(14)*. In conclusion, we present evidence for immune maturation as well as maintenance of potentially pathogenic autoreactive memory and plasma cells in the rheumatic joint already at the onset of the disease.

Comparing the findings of Zhang et al. *(8)* with our study in more detail, we could not find a separate cluster with autoimmune-associated CD11c+ B cells. This may be a more prominent feature of established RA, i.e. a consequence of chronic inflammation or immunosuppressive therapy.

While our approach to combine single cell and spatial transcriptomics for two ACPA- and two ACPA+ samples gives a very detailed picture, the small sample size is limiting, when drawing conclusions about the disease.

Intriguingly, we found a new plasma cell derived ACPA that displays some of the classic features previously attributed to RA autoantibodies such as N-linked glycosylation sites in the Fab fragment introduced by SHM *(15–17)*. However, this new tissue plasma cell-derived ACPA appears unique with a narrow pattern of reactivity compared to previously described ACPA *(18– 23)*. The A4802:3286 ACPA clone reacted exclusively and strongly to a mutated vimentin peptide originally identified in RA joint tissue *(24)*.

60% of our plasma cell derived monoclonal antibodies originated from expanded clones with representatives in the memory and plasma cell compartment. Presence of clonally expanded B cells has been associated with increased likelihood of RA onset in at risk individuals *(9, 25)*. However, while our data suggests a local B cell differentiation process, egression from the circulation may be an alternative explanation for the observation of clonal expansions in the joints.

Furthermore, we found six polyreactive plasma cell clones. Polyreactivity among mutated monoclonal antibodies has been reported in many different settings e.g. HIV infection and vaccination *(26, 27)*. It has previously been demonstrated that there is a pool of polyreactive switched memory B cells that could originate from bypassing a tolerance checkpoint even in healthy donors *(28)*. We speculate that differentiation of these polyreacitve switched memory B cells into plasma cells perpetuates RA locally and systemically.

Our study included synovial biopsies from untreated RA patients sampled within two days of diagnosis, hence the transcriptomic data we report are not biased by treatments such as steroids and disease-modifying anti-rheumatic drugs that most RA patients receive. We find many similarities to other data sets analyzed much later in the disease process demonstrating that immune pathways implicated in disease pathogenesis are at play already at this early time point. We also demonstrate that high throughput technologies can be utilized to generate monoclonal antibodies from patient-derived B cells. Amongst the plasma cells in the inflamed joint, we identified an ACPA with strong reactivity to a modified citrullinated vimentin peptide, a specificity previously linked with osteoclast activation *(22)*. Future studies will show if this new ACPA also has functional capacities.

## MATERIALS AND METHODS

### Study Design

In this cross-sectional study, we collected synovial tissue from eight Rheumatoid Arthritis (RA) patients. The inclusion criteria were that the biopsies were taken at the time of diagnosis and that the patients were naïve to anti-rheumatic treatment. Since RA starts with manifestations in small joints, we initially sampled small joints such as the metacarpophalangeal joint. However, due to low B cell recovery from this site, we instead focused on sampling larger joints such as wrists and knees. Sorted B cells from inflamed joints of five RA patients were submitted for scRNAseq. The researchers performing the experiments were aware of the clinical diagnosis of the patients. Sample numbers are provided in figure legends.

### Patients

All RA patients were recruited from the Rheumatology Clinic of the Karolinska University Hospital. All biopsies were performed in accordance with the Helsinki Declaration and written informed consent was given by each patient before entering the study. Clinical features of the patients are presented in Supplemental Table 1. We selected three patients that had high quality scRNAseq data and one additional patient for the spatial transcriptomics. These four RA patients were female, 50-75 years old and had high disease activity.

### Sample processing

Arthroscopic biopsies were taken from large joints and ultra-sound guided biopsies from small joints of RA patients within two days of diagnosis.

One set of the biopsies were collected for scRNAseq in cold RPMI medium (Gibco) supplemented with 1% penicillin streptomycin and 1% glutamine. The tissue was digested in 4mg/ml Collagenase A (Roche) and 0.1mg/ml DNAse (Roche) in RPMI medium at 37°C while shaking. After 1h, ice cold RPMI medium supplemented with 10% FBS was added to stop the enzymatic reaction. Further descriptions of the staining are in supplementary materials and methods. Cells were sorted on a Becton Dickinson influx cell sorter. The gating strategy is depicted in Supplementary Figure 1. 3,468 single cells were sorted into 384-well plates containing lysis buffer, before snap frozen. We then submitted plates with ≥382 cells/donor for scRNAseq reducing our patient pool to five.

Another set of tissue pieces were snap frozen in isopentane prechilled with liquid nitrogen and stored at -70°C for immunohistochemistry and 10x Visium analyses which are described in the supplementary materials and methods section. Notably, the sections for the different analyses were 50-100µm apart, so some variance in tissue morphology could be expected.

### Single cell library preparation, sequencing and analysis

Single cell libraries were prepared according to the SmartSeq2 protocol *(29)*. A brief description is in the supplementary materials and methods. Libraries were pooled per plate and sequenced on an Illumina NovaSeq with 2×100bp paired-end reads yielding a median of 2.5M reads/well. Each plate had two empty control wells. Flow cytometry and scRNAseq data analysis are in the supplementary materials and methods.

### 10x Visium data analysis

The spatial gene expression data was analyzed with the STUtility package *(30)* and the Seurat package v.3.2.2 *(31, 32)*. Each section was individually normalized with the SCTransform function which uses a regularized negative binomial regression to transform the UMI count data. Thereafter donor integration was performed with the Harmony algorithm followed by a Shared Nearest Neighbor (SNN) construct graph and clustering using default settings. The clusters were colored and the harmony embedded data was visualized in an UMAP and on the tissue sections. The raw counts of the clustered data underwent a differential gene expression analysis with logFC>1 and adjusted p-value<0.05 cut-offs to define sets of upregulated genes.

The factor analysis was run with a Non-negative Matrix Factorization specifying 20 factors on all batch corrected SCTransformed sections. The pathway analysis was performed with the BioPlanet 2019 tool *(33)* on the top-20 genes from the factor analysis.

### Single cell and spatial data integration

Significantly enriched genes and overlapping gene signatures were identified according to the supplementary materials and methods. The single cell data from Zhang et al. *(8)* was predicted on the spatial tissue data with the FindTransferAnchors and TransferData functions from the Seurat package with default settings. This was performed with SCTransformed data on both the single cell data set and the spatial data set. Spearman correlation was used as an estimate for co-localization of the SC-B2 and SC-B4 cells against all other single cell types. Besides, we showed raw counts for *CXCL12* and *VIM* expression, respectively.

### B cell receptor reconstruction and expression

For B cell receptor reconstruction from the single cell data, we used BraCeR *(34)*. A brief description of filtering and visualization can be found in the supplementary materials and methods. Selected immunoglobulin heavy and light chain pairs were cloned into expression vectors *(35)*. Recombinant monoclonal antibodies were expressed in the Expi293 system (Thermo Fisher Scientific). Purified antibody concentration was determined by ELISA.

### Reactivity testing

Monoclonal antibody binding was investigated at 5µg/ml. Anti-CCP2 ELISA (Euro Diagnostica) was performed according to manufacturer’s instructions. The serum (1:50 dilution) cut-off for positivity was 25units/ml. For the polyreactivity ELISA, soluble membrane protein (SMP) extract was coated at 5µg/ml *(35)*. The ACPA was further characterized by a modified Vimentin peptide ELISA (Orgentec Diagnostika) containing citrulline, acetyl-lysine, homocitrulline, acetyl-ornithine or ornithine residues. The ACPA fine specificity was further evaluated using a custom-made peptide array (Thermo Fisher Scientific, ImmunoDiagnostics) containing citrullinated peptides and control antigens *(36)*.

## Data Availability

Raw sequencing data, count matrices, cluster labels and BCR sequences are available through an MTA upon request.

## Acknowledgements

We thank Heidi Wähämaa and Marianne Engström for help with collecting the biopsy material, Sara Turcinov and Vijay Joshua for guidance on experimental setups, Caroline Grönwall for organizing antibody cloning, Ragnhild Stålesen for help with antibody expression, Holger Bang for providing the Orgentec plates and Monika Hansson for investigating antibodies on the citrullinated peptide array. We equally thank Fan Zhang and Soumya Raychaudhuri for sharing of their published data *(8)*. We also acknowledge the Eukaryotic Single Cell Genomics facility (ESCG) at SciLifeLab, Stockholm, for preparing sequencing libraries. The authors acknowledge support from the National Genomics Infrastructure in Stockholm funded by Science for Life Laboratory, the Knut and Alice Wallenberg Foundation and the Swedish Research Council, and SNIC/Uppsala Multidisciplinary Center for Advanced Computational Science for assistance with massively parallel sequencing and access to the UPPMAX/SNIC-SENS computational infrastructure partially funded by the Swedish Research Council through grant agreement no. 2018-05973.

## Author contributions

Conceptualization: UH, KC, PLS, VM

Experimentation: UH, KC, EK, PS, AV, SHM, LI, KA, MK

Data analyses: UH, KC, PS, LL, SHM, LI, KC, MK

Interpretation: UH, KC, KC, MK, GBKH, AIC, SAT, PLS, VM

Writing: UH, KC, VM

## Data and materials availability

Raw sequencing data is available on the European Genome-Phenome Archive under the accession number xyz. Count matrices, cluster labels and BCR sequences are available through an MTA upon request.

