## Supplementary materials for "Integrated Single Cell and Spatial Transcriptomics Reveal Autoreactive Differentiated B Cells in Joints of Early Rheumatoid Arthritis"

**Supplementary Materials and Methods**

Flow cytometry staining

The tissue was strained through a 40µm mesh. Cells were washed and counted in ice cold PBS (Gibco). Cells were resuspended in 50µl PBS and stained in antibody staining mixture containing 3µl anti-CD19 BV421 (HIB19), 10µl anti-CD3 FITC (UCHT1), 10µl anti-CD14 FITC (M5E2), 5µl anti-IgD BV605 (IA6-2), 5µl anti-CD138 APC (MI15), 5µl anti-CD27 PE-Cy7 (M-T271, all Becton Dickinson) and 12µl 1% human serum for 30min at 4°C. Cells were washed, resuspended in 50µl AnnexinV binding buffer and stained with 4µl AnnexinV FITC for 10min at RT. Finally, cells were diluted in AnnexinV binding buffer.

Immunohistochemistry

Cryostat sections (7µm) were fixed with 2% formaldehyde (Sigma) for 20min. Sections were incubated with mouse anti-CD19 (HD37) mixed with mouse anti-CD20 (L26) antibodies from Dako Denmark A/S for validation of B cells according to a described protocol *(37)*. Isotype matched irrelevant antibodies were used as a negative control. The staining sections were examined using a Polyvar II microscope (Reichert-Jung, Vienna, Austria) and Leica Qwin IM500 software (Leica, Cambridge, UK).

Spatial RIN analysis

The Spatial RIN assay was used to assess the quality distribution of RNA over the biopsies. The protocol was previously described *(38)*. The sectioning thickness was set to 7µm. Fixation was performed in 100% methanol in freezing conditions for 30min and after staining and imaging, an

8min permeabilization followed. The tissue removal step was performed for 1h in 56°C with interval shake using 4x β-mercaptoethanol diluted in Buffer RLT Plus (QIAGEN). Further steps were performed as described.

10x Visium processing

The Visium Spatial Tissue Optimization Slide & Reagent kit (10x Genomics) was used to optimize permeabilization conditions for the fresh frozen tissue biopsies. The manufacturer's protocol was followed and the optimal permeabilization time was determined to be 15min.

A total of ten sections divided into four patient samples were sectioned with 7μm thickness and mounted onto Visium Spatial Gene Expression Slides & Reagent kits (10x Genomics). Sequencing libraries were prepared following the manufacturer's protocol, with permeabilization performed for 15min, as established in the TO assay. The 10x Visium spatial transcriptomics libraries were prepared and sequenced on an Illumina NextSeq with the 150 cycles high output kit. The libraries were sequenced with 28bp from Read1 and 120bp from Read2, and at a depth of about 100M reads/sample.

10x Visium data processing

Following demultiplexing of BCL files, the Read2 FASTQ files were trimmed using Cutadapt *(39)* to remove full-length or truncated template switch oligo (TSO) sequences (AAGCAGTGGTATCAACGCAGAGTACATGGG) from the 5' end and polyA homopolymers from the 3' end. We required partial matches up to 5bp or intact TSO sequences with a maximum of 3 errors. For the 30 homopolymer trimming, a sequence of 10 A's was used regardless of the position in the read. The trimmed data were processed with the Space Ranger pipeline (version 1.0.0, 10x Genomics) and mapped to the GRCh38 v93 genome assembly.

ScRNAseq library preparation

In brief, mRNA from single cells and ERCC spike-in were reverse transcribed using an Oligo(dT) primer, before a locked nucleic acid strand switch primer was added. cDNA amplification was primed by ISPCR primers and yields were assessed on an Agilent Bioanalyzer. The PCR product was tagmented using a Tn5 transposase integrating Read1 and Read2 sequences at the termini of the target DNA. Libraries were indexed using the Nextera XT DNA indices and assessed for concentration on the Agilent Bioanalyzer.

Flow cytometry and scRNAseq data analysis

FCS files were analyzed using FlowJo and R. Single cell data was extracted using InfluxIndex2CSV.

Raw reads were processed using FastQC. Adapter sequences were trimmed using TrimGalore and mapped using STAR. Low quality cells that resembled empty control wells in terms of quality metrics were excluded by assessing a threshold along the first principal component vector in R. The quality metrics included the Q30 score, the number of raw read sequences, the number of failed FastQC metrics, before and after trimming, the number and length of mapped reads, mapped genes and ERCC reads.

The ERCC spike-ins were labelled by the scran::isSpike function. We computed technical and biological size factors by using the scran::computeSpikeFactors and scran::computeSumFactors functions. We normalized the raw read counts of endogenous genes with the biological size factors, and that of spike-ins with the technical size factors using the scater::normalize function. We fitted a curve to the mean-variance values of spike-ins using the scran::trendVar function. The fitted value is used to estimate the technical component of the total variance of an endogenous gene. We used the scran::decomposeVar function to find genes with significantly higher variation than the fitted technical variation (biological variance>0.5 and FDR $\leq 0.05$). These 2747 highly variable genes were kept for UMAP dimensionality reduction using the scater::runUMAP function. Three clusters were visually identified. For cluster labeling, we used a k-means clustering identifying 5 clusters. One cluster was composed of naïve B cells, one cluster was composed of plasma cells and three clusters were joined and composed of memory cells. For differential gene expression analysis, we selected the top-20 differentially expressed genes by assessing the average log2FC of expression for each cluster against the other two.

Single cell and spatial data integration

To find significantly enriched genes for memory B cells and plasma cells from our data, and SC-B1, SC-B2 and SC-B4 from the Zhang et al. study *(8)*, we used Kruskal-Wallis Test with Dunn correction and performed p-value adjustment for multiple testing according to the Bonferroni method. As comparators, we only used the previously mentioned clusters as well as the cluster of naïve B cells from our data. Significantly enriched genes were selected based on average expression FC>1 and adjusted p-value<0.01, whilst the comparators were ignored. To find overlapping gene signatures of the clusters from both datasets, we used Fisher's exact test under the $H_0$ hypothesis that there are random associations between the significantly enriched B cell markers in both data sets.

B cell receptor filtering and visualization

The output tables from BraCeR *(34)* were filtered based on completeness and expression levels of the paired V(D)J transcripts using a customized R script. Combined heavy and light chain sequences were aligned using Clustal Omega. The alignment was visualized using FigTree.

### Supplementary Figures

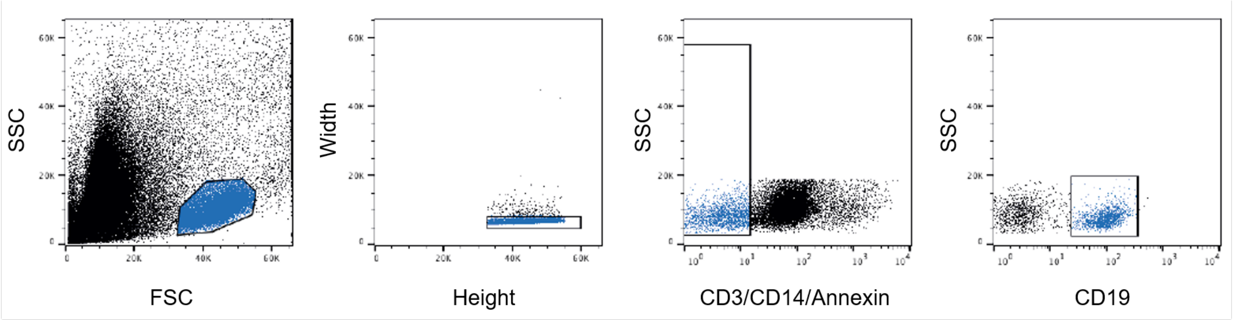


Fig. S1. The gating strategy for the B cell sort is shown. We selected AnnexinV-CD3-CD14-CD19+ single lymphocytes.

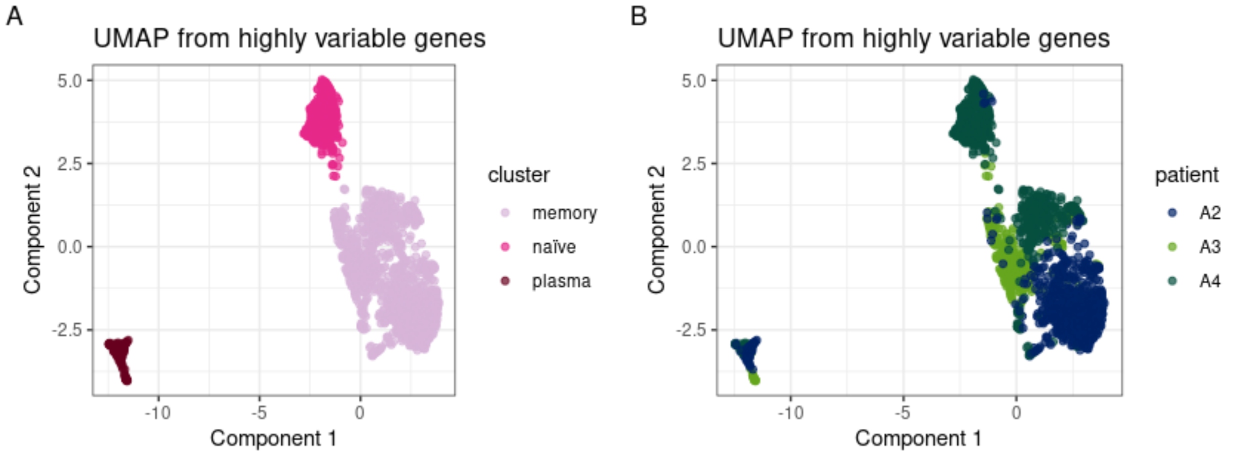


Fig. S2. UMAP of scRNAseq data coloured by **(A)** cell cluster and **(B)** patient contribution. Notably, the plasma cell cluster was the most distinct, while the naïve and memory cell clusters were more similar. The plasma cell and memory B cell clusters had contributions from all three patients, while the naïve B cell cluster was mainly (95%) composed of cells from one of the ACPA- RA samples (A4). This is consistent with the large naïve B cell population (42%) observed for this patient in the flow cytometry data. Also, the memory cell cluster was influenced by patient specificity

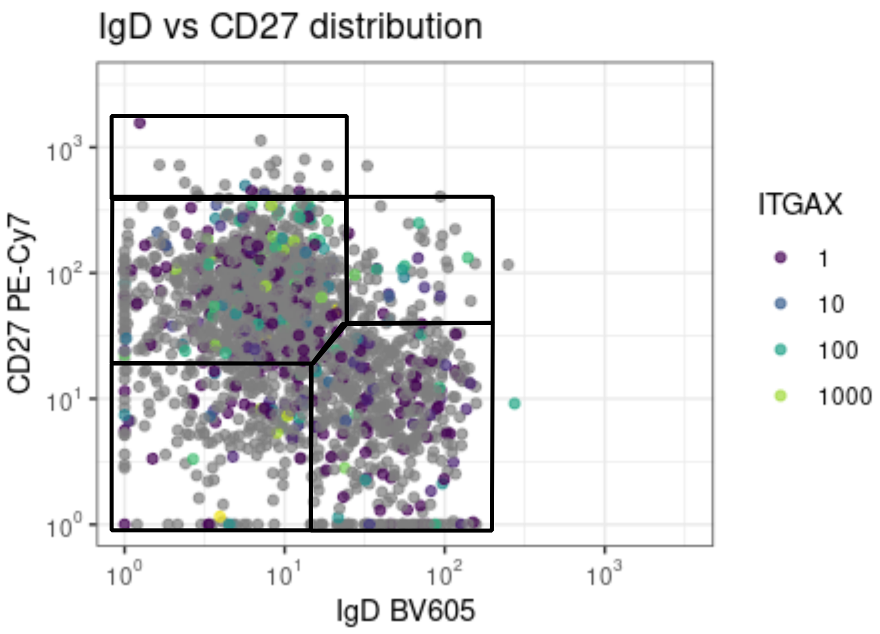


Fig. S3. ITGAX (encoding CD11c) expression was projected onto the IgD vs CD27 distribution. We did not find enrichment of *ITGAX* in the double negative population, which would indicate autoimmunity-associated B cells.

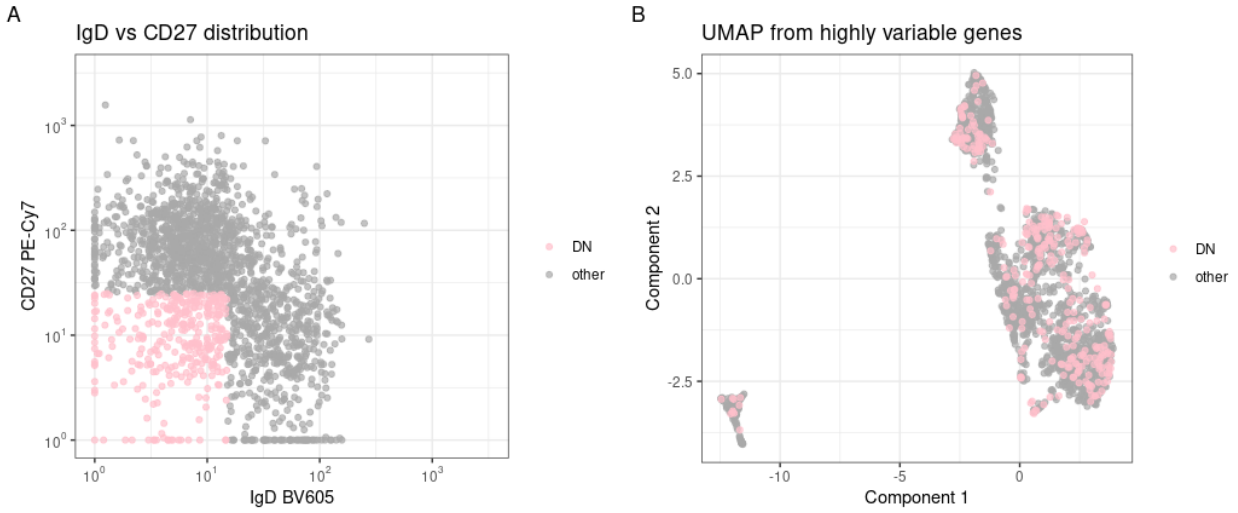


Fig. S4. The double negative population in the scRNAseq data is shown. Cells defined by the double negative gate in the flow cytometric analysis mapped to all clusters in the single cell transcriptomic data.

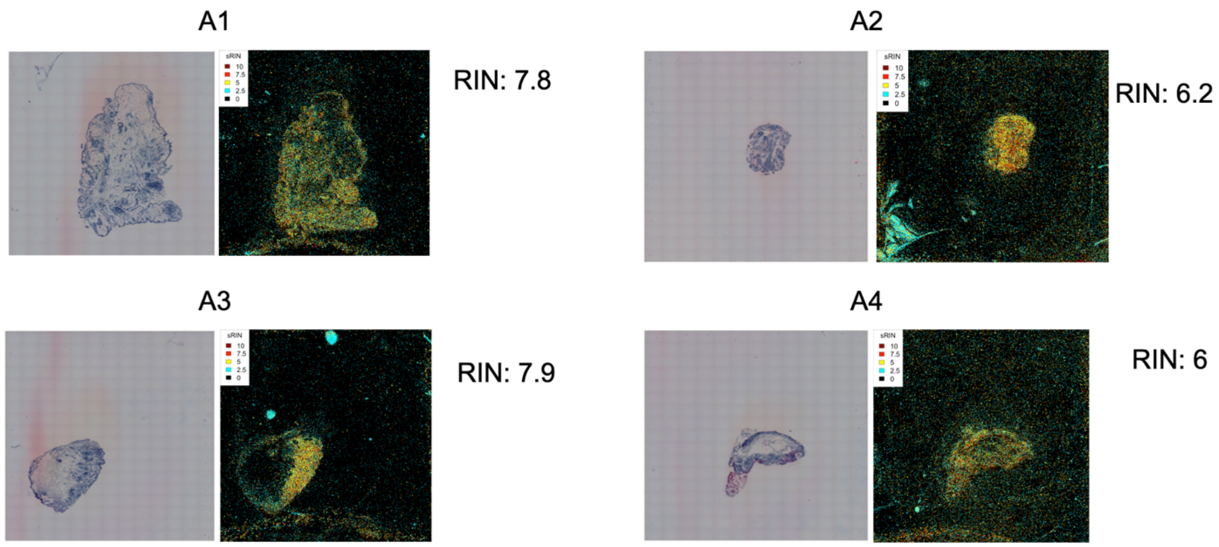


Fig. S5. Spatial RIN. Heatmap over the spatial RIN scores with bulk RIN values from extracted material shown next to the heatmaps

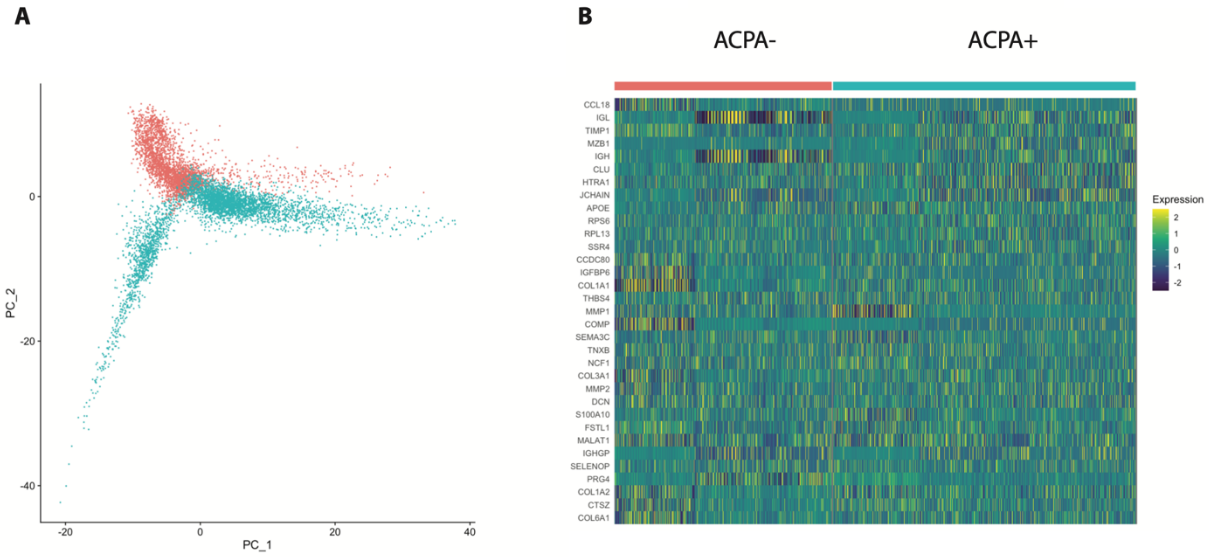


Fig. S6. **(A)** PCA labelled by serotype (red ACPA-, turquoise ACPA+) showing overlaps and separation between data points for each group. **(B)** Heatmap over the marker genes for each group with little differences between ACPA+ and ACPA- RA.

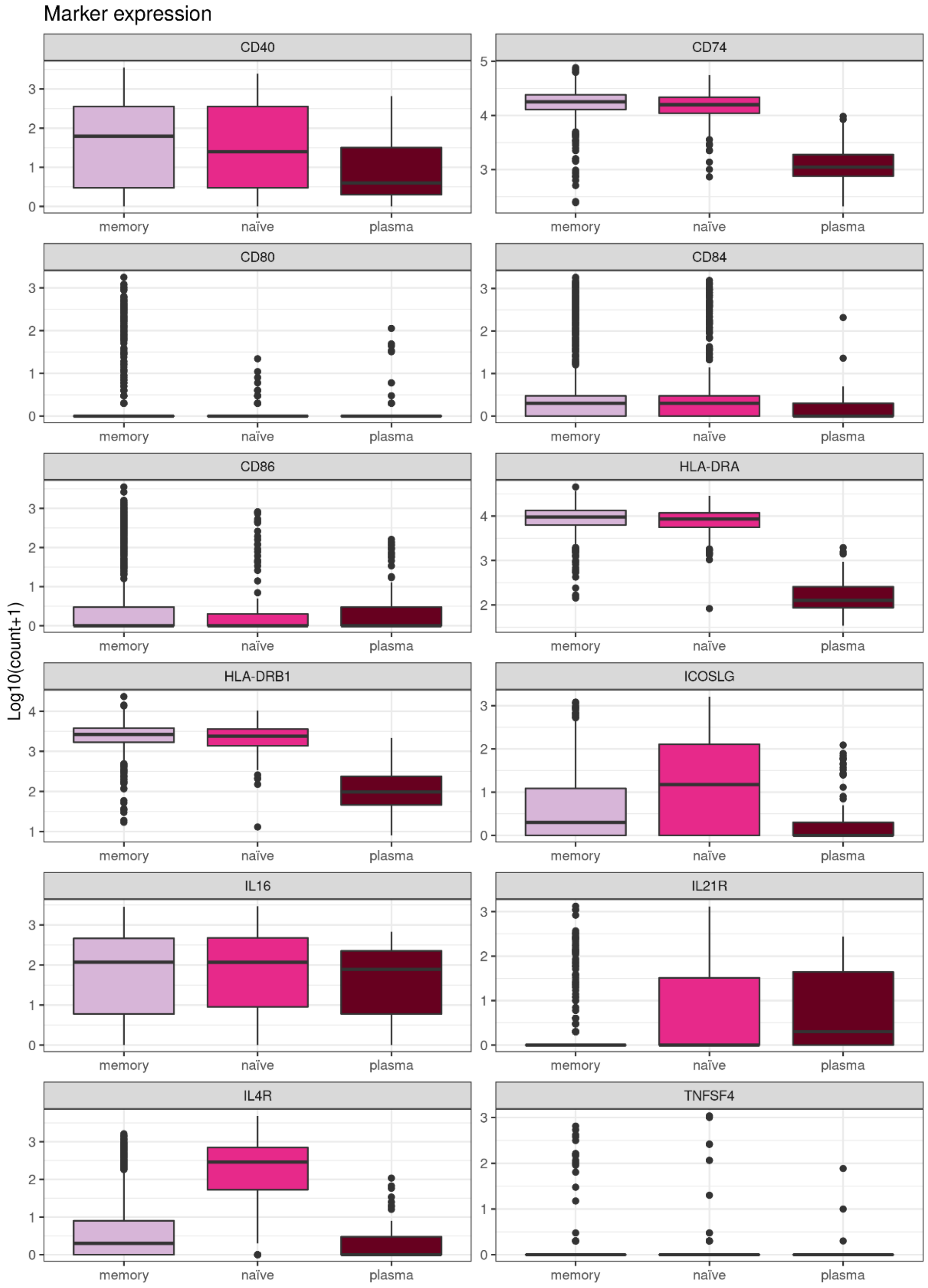
Marker expression
CD40
CD74
CD80
CD84
CD86
HLA-DRA
HLA-DRB1
ICOSLG
IL16
IL21R
IL4R
TNFSF4
memory
naïve
plasma
Log10(count+1)

Fig. S7. Logarithmic read count for B-T cell interaction markers are shown grouped by cell subset. Most markers are present in the naïve and the memory cell subset.

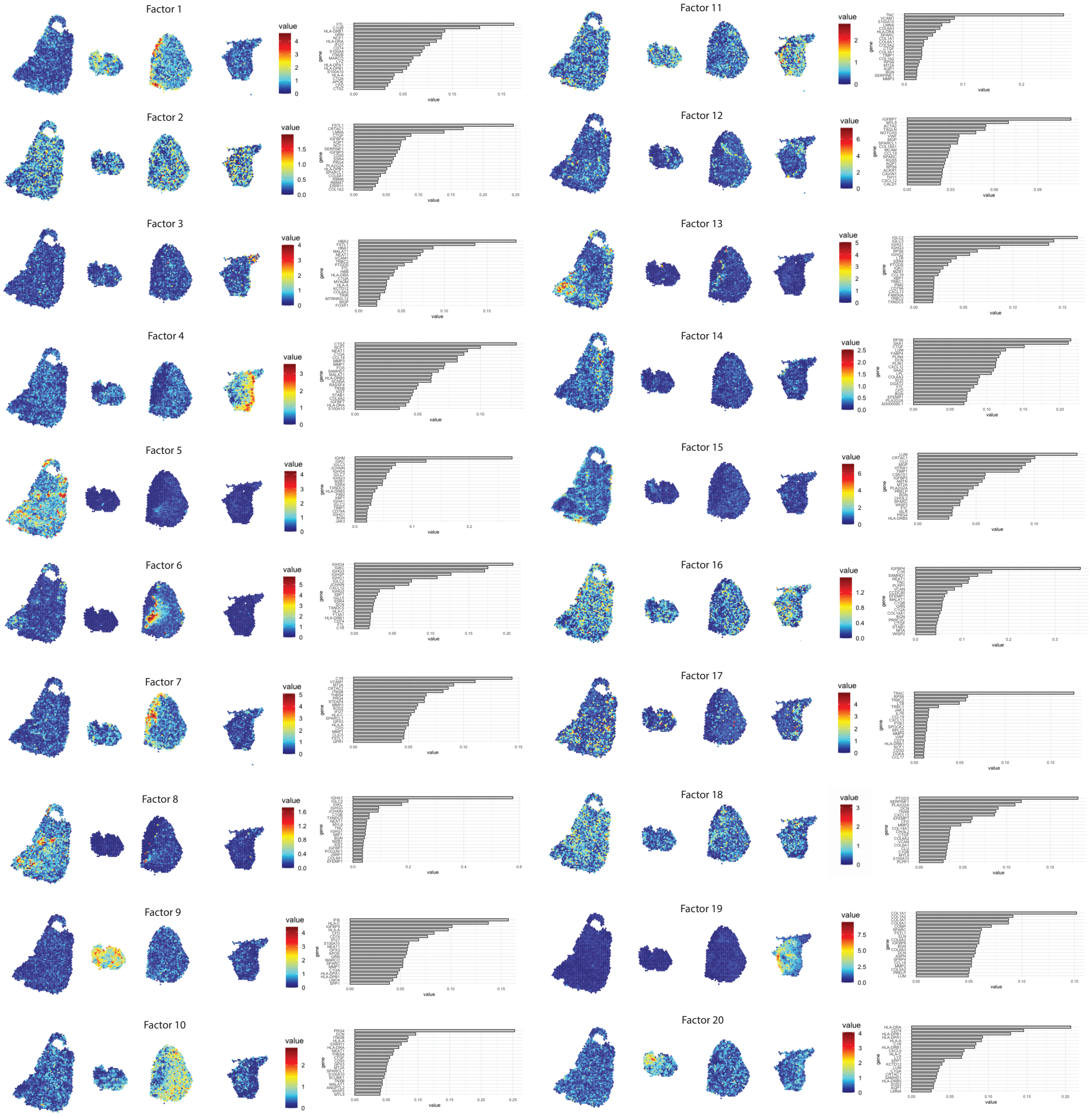


Fig. S8. The factor analysis for spatial transcriptomics is shown. Most factors (e.g. 2,11,16,18) are represented on all tissues. Several factors have expression activities overlapping with distinct morphological areas such as infiltrates, connective tissue as well as areas bordering the infiltrates. These could be regarded as baseline tissue signals with *CD55* being of particular interest as it marks the intimal lining fibroblast-like synoviocytes. Three factors (6,8,13) shows a strong activity towards the immune cell-rich areas in biopsy A1. These areas correlate with IGH, IGL, *JCHAIN*, *MZB1*, *XBP1* and *CD79A* expression suggesting plasma cell presence. Factor 20 has strong activity within cluster 4 for biopsy A2. Here the driver genes are the plasma cell markers mentioned before as well as *HLA-DRB1* and *CD74* further highlighting antigen presentation. Biopsy A3 present factor 6 with strong activity in lymphocyte aggregates. Factor 6 is among others associated with the HLA-associated *CD74* gene. For biopsy A4, two of the factors (4,19) have genes associated with connective tissue and several HLA genes.

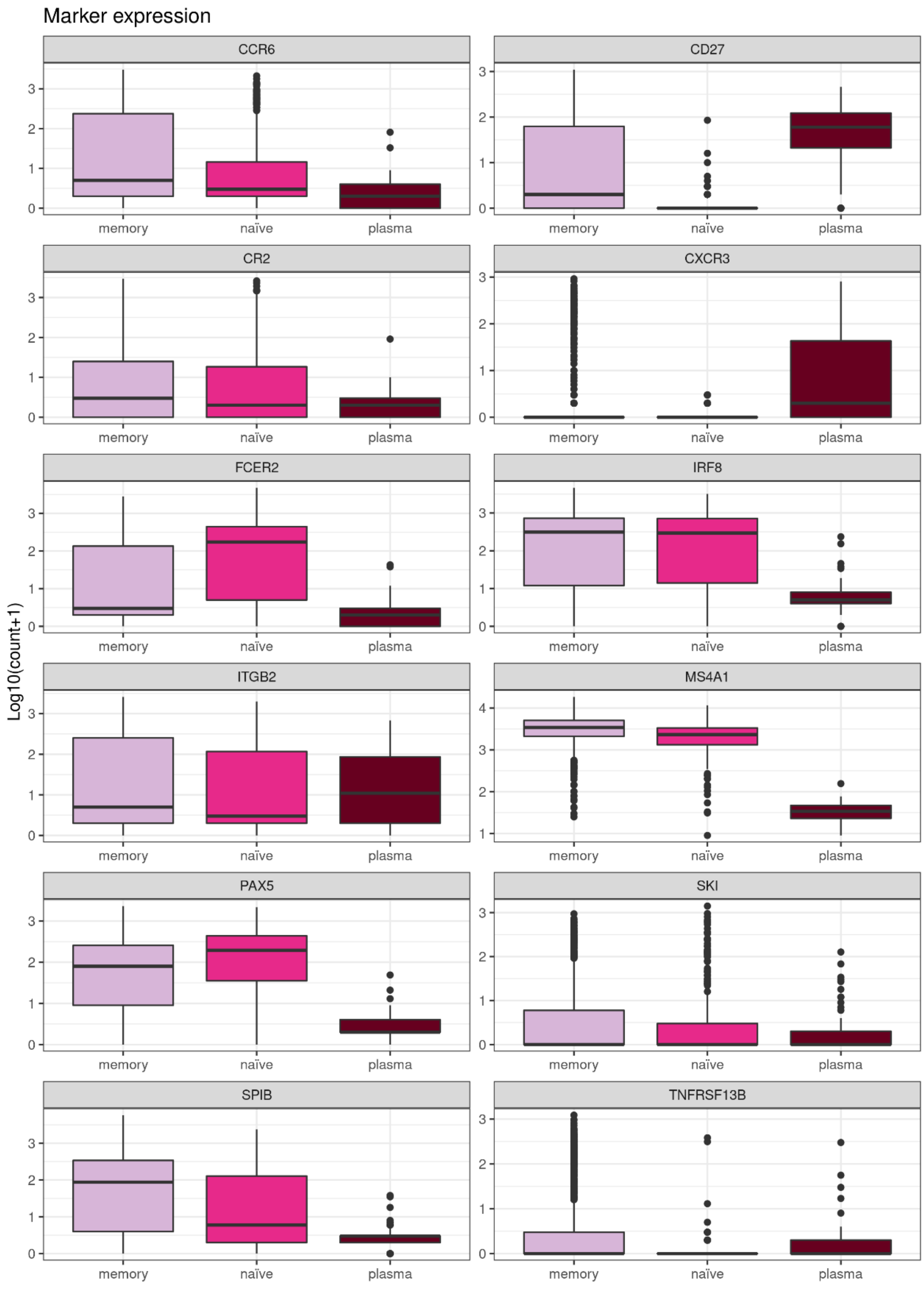


Fig. S9. Logarithmic read count for memory B cell markers is shown grouped by cell subset.

Most memory markers are either shared with the naïve or the plasma cell subset.

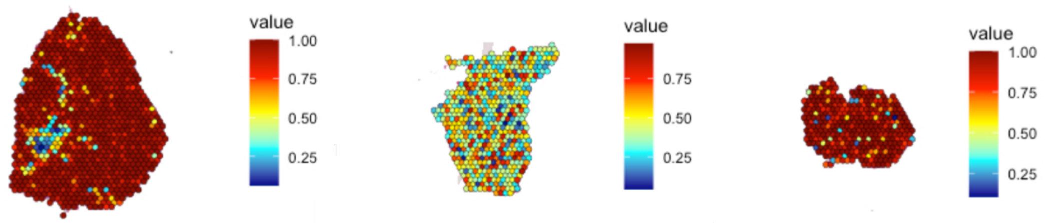


Fig. S10. The memory B cell subset from paired data from the same individual was predicted on the tissue sections.

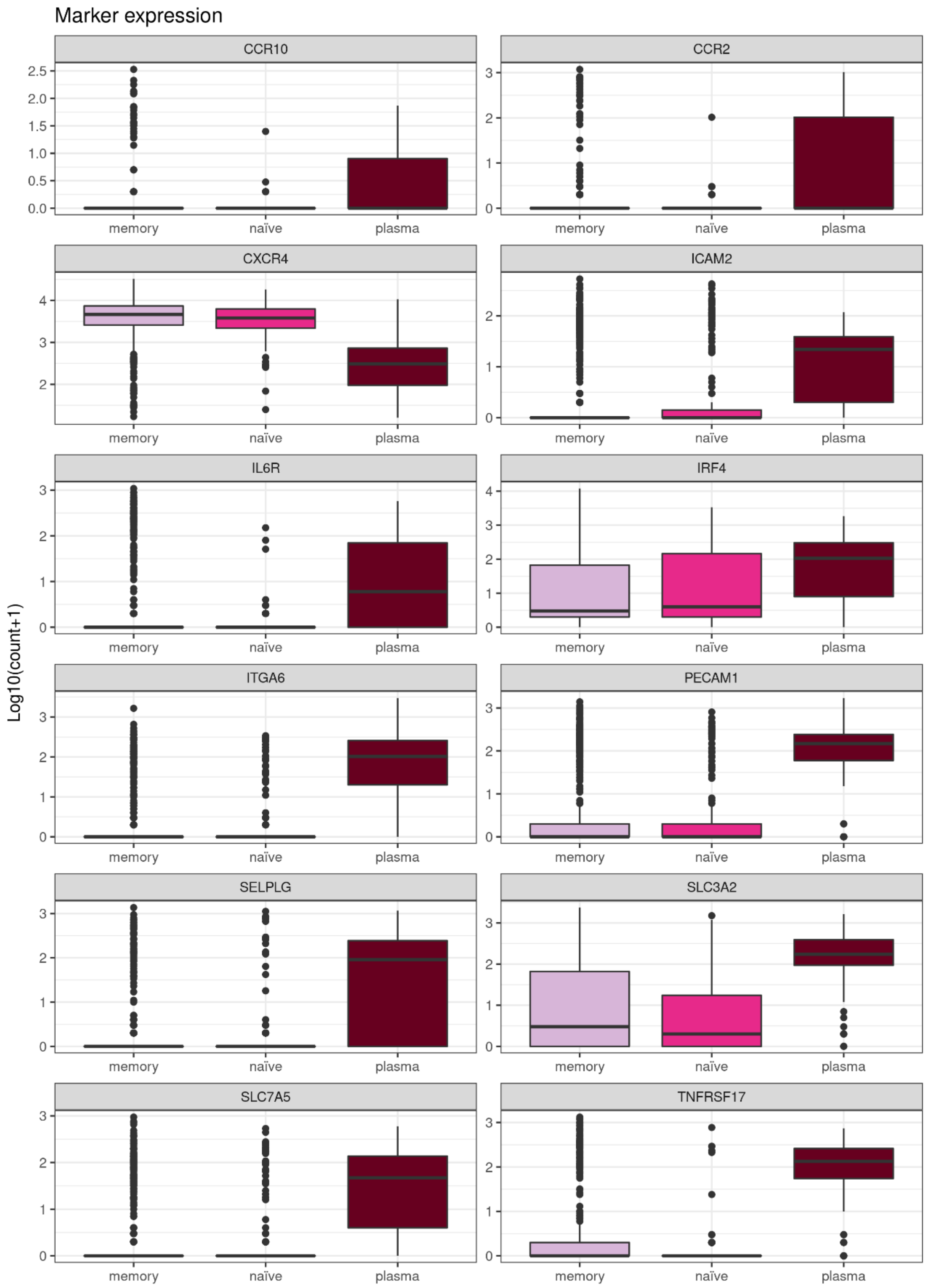


Fig. S11. Logarithmic read count for plasma cell markers is shown grouped by cell subset. Most markers are clearly exclusive to plasma cells.

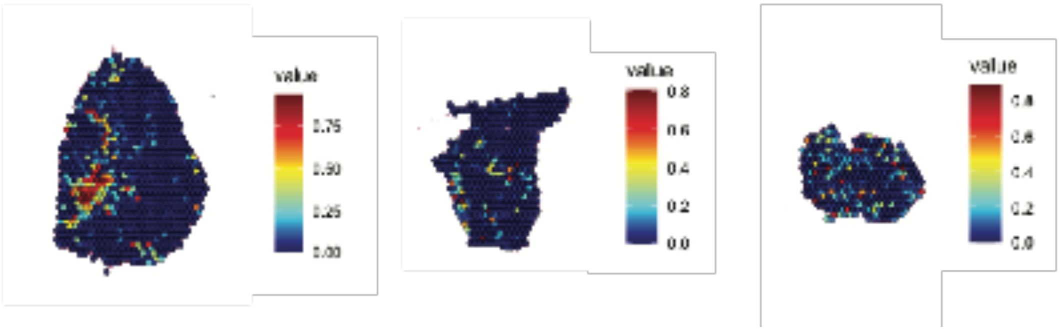


Fig. S12. The plasma cell subset from paired data from one individual was predicted on the tissue sections. Integration showed a clear correlation with the Zhang et al. *(8)* data.

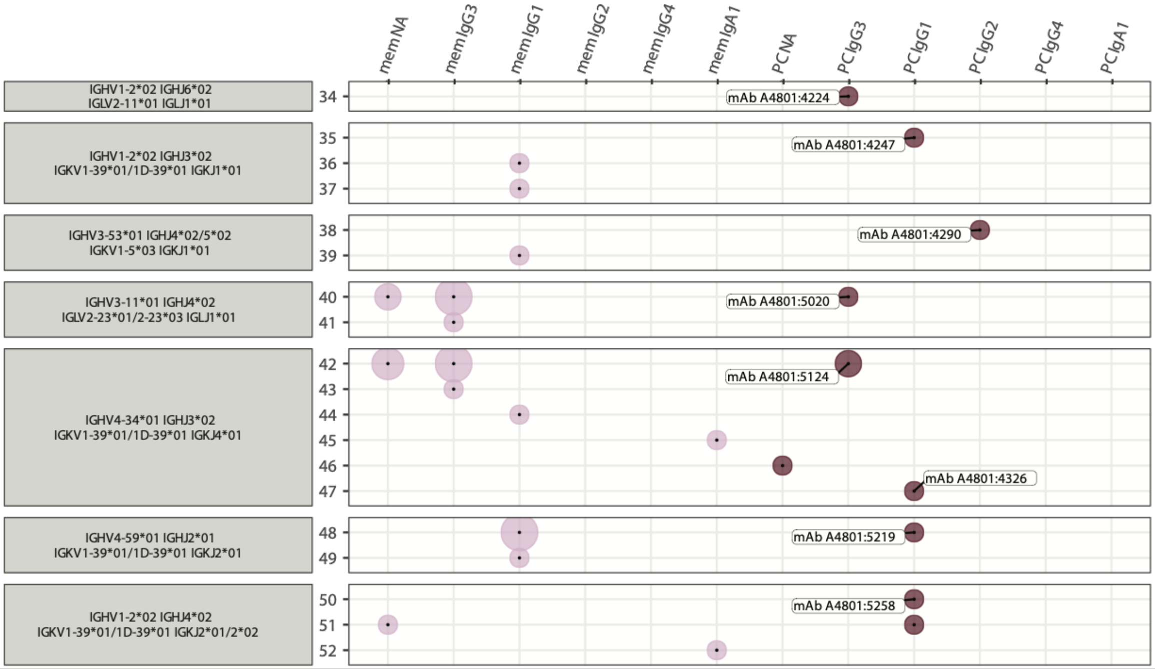


| Clone | HCDR3 | VH SHM (%) | LCDR3 | VL SHM (%) |
|---|---|---|---|---|
| 34 | AGTNYDFWSGRPWYYYGMDV | 1/294 (0.3) | CSYAGSYTYV | 0/295 (0.0) |
| 35 | VRAARGSGWFHDVFDI | 6/295 (2.0) | QQSYSTPRT | 3/284 (1.1) |
| 36 | ARANFGGLRGYDAFDI | 6/295 (2.0) | QQSYSTRT | 3/283 (1.1) |
| 37 | ARANFGGLRGYDAFDI | 8/295 (2.7) | QQSYSTRT | 2/283 (0.7) |
| 38 | AKDGSSRSLVS | 10/293 (3.4) | QQYNSFPWT | 11/280 (3.9) |
| 39 | AKDGSSRSLVS | 15/293 (5.1) | QQYNSYSPTWT | 1/287 (0.3) |
| 40 | ARDKRRRHYDSSGYHFDY | 6/296 (2.0) | CSYAGSSTSLYV | 1/295 (0.3) |
| 41 | ARDKRRRHYDSSGYHFDY | 6/295 (2.0) | CSYAGSSTSLYV | 1/295 (0.3) |
| 42 | ARARSGISAIHGVFDI | 3/293 (1.0) | QQSYSTPPT | 4/287 (1.4) |
| 43 | ARARSGISAIHGVFDI | 3/285 (1.1) | QQSYSTPPT | 4/287 (1.4) |
| 44 | ARARSGLGATHGAFDI | 7/292 (2.4) | QQSYSTPPT | 1/287 (0.3) |
| 45 | ARGGGYGGNSVRRAFDI | 28/293 (9.6) | QQTYITPRA | 21/284 (7.4) |
| 46 | ARARSGLGATHGAFDI | 4/292 (1.4) | QQSYSTPPT | 4/287 (1.4) |
| 47 | ARARSGLGATHGAFDI | 6/292 (2.1) | QQSFSTPPT | 4/287 (1.4) |
| 48 | ARGKTGYSSRNHWYFDL | 2/292 (0.7) | QQSSSTPMYT | 2/284 (0.7) |
| 49 | ARGKTGYSSRNHWYFDL | 2/292 (0.7) | QQSYSTPLST | 2/284 (0.7) |
| 50 | ARGIMGQFPPDY | 2/293 (0.7) | QQSYSTPQT | 4/286 (1.4) |
| 51 | ARGIVGQFPPDH | 8/293 (2.7) | QQSYSTPQT | 2/286 (0.7) |
| 52 | AREVGVGAILDY | 30/296 (10.1) | QQSYSTPYT | 15/284 (5.3) |

Fig. S13. The plot shows the equivalent clonotype information that is shown in figure 6A for the seven plasma cell derived monoclonal antibodies originating from A3.

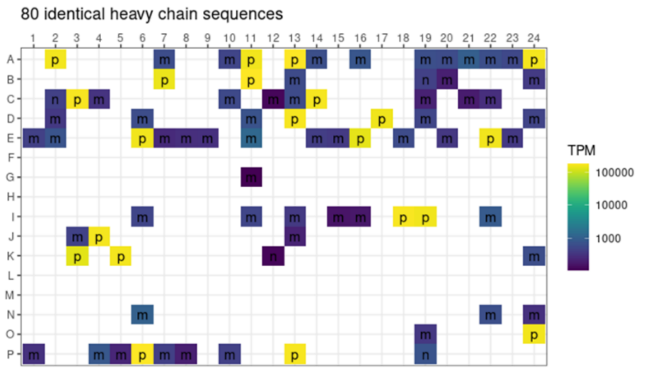


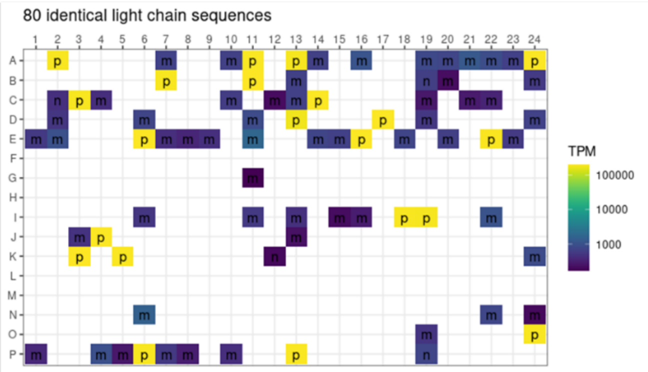


Fig. S14. No index-hopping for monoclonal antibody A4797:2017. Cells expressing the A4797:2017 antibody were derived from plasma cells (p), memory B cells (m) and naïve B cells (n).

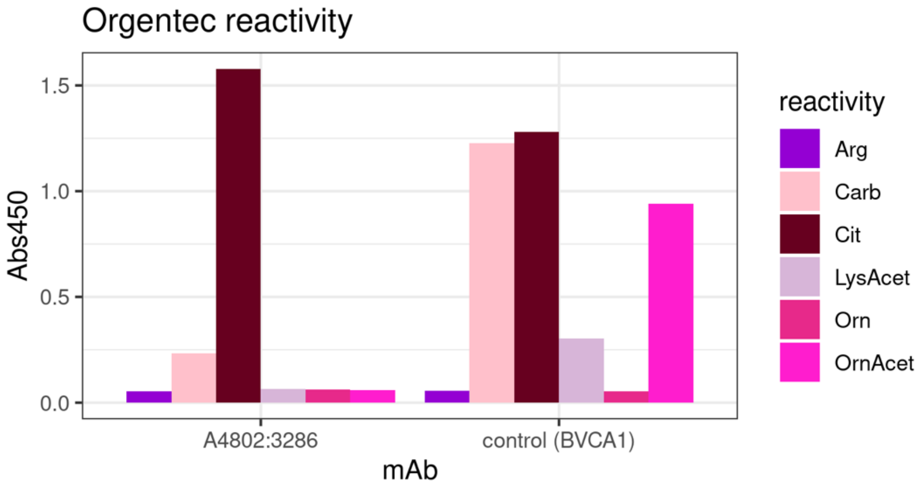


Fig. S15. The Orgentec reactivity of the A4802:3286 clone compared to control (BVCA1 *(16)*) is shown. The ACPA clone was found to be negative for the acetylated peptide, weakly positive for the carbamylated peptide but strongly positive for the citrullinated peptide.

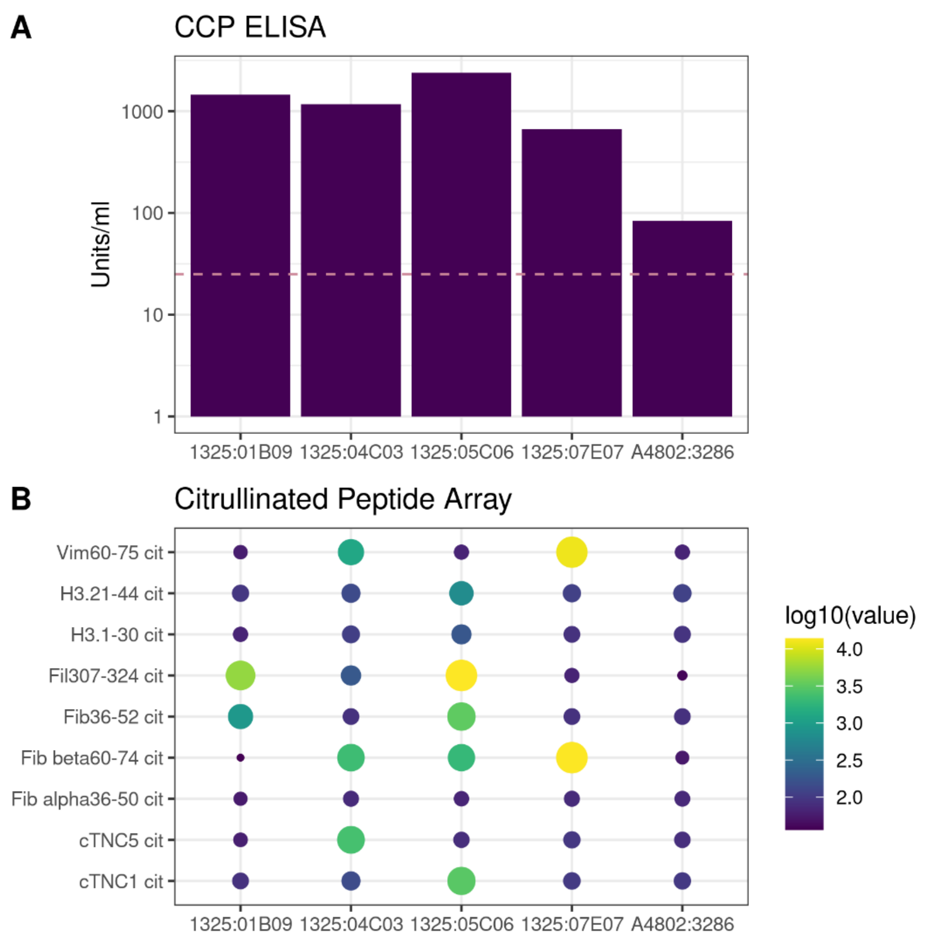


Fig. S16. **(A)** CCP2 ELISA and **(B)** citrullinated peptide array *(36)* to test antigen specificity. We compared the identified A4802:3286 ACPA with previously described ACPAs generated from plasma cells from synovial fluid of RA patients with long lasting disease *(22)*. The cut-off for the anti-CCP2 ELISA was 25units/ml. On the citrullinated peptide array, we assessed reactivity towards citrullinated Vimentin (Vim), citrullinated Histone 3 (H3), citrullinated Filaggrin (Fil), citrullinated Fibrinogen (Fib) and citrullinated Tenascin-C (TNC). Numbers indicate amino acid residues.

| mAb | N-X-S/T glycosylation sites germline VH | VH | germline VL | VL |
|---|---|---|---|---|
| A4797:2017 | 0 | 0 | 0 | 0 |
| A4797:2216 | 0 | 0 | 0 | 0 |
| A4801:4224 | 0 | 0 | 0 | 0 |
| A4801:4247 | 0 | 1 | 0 | 0 |
| A4801:4290 | 0 | 0 | 0 | 0 |
| A4801:4326 | 2 | 2 | 0 | 0 |
| A4801:5020 | 0 | 0 | 0 | 0 |
| A4801:5124 | 1 | 1 | 0 | 0 |
| A4801:5219 | 0 | 0 | 0 | 0 |
| A4801:5258 | 0 | 0 | 0 | 0 |
| A4802:3086 | 0 | 0 | 0 | 0 |
| A4802:3286 (ACPA) | 0 | 0 | 0 | 2 |
| A4802:3314 | 0 | 0 | 0 | 0 |
| A4802:6090 | 0 | 1 | 0 | 1 |
| A4802:6117 | 0 | 0 | 0 | 0 |

Fig. S17. Germline and germline-diverted glycosylation sites in monoclonal antibodies are shown.

**Supplementary tables**

| Diagnosis | subjects N, gender | age median (range) | CCP titre (cut-off 3) | RF status + / - | disease activity DAS28-CRP | symptom duration months |
|---|---|---|---|---|---|---|
| ACPA+ RA | 4F | 58.5 (23-74) | 80 - 300 | 3/1 | 4.93 (3.85-6.66) | 5.75 (2-24) |
| ACPA- RA | 3F/ 1M | 75.5 (67-80) | <3 | 1/3 | 6.10 (5.50-6.40) | 4.5 (3.5-5) |

Table S1. Clinical parameters of studied patients are shown.

| cluster | gene | p_val | avg_logFC | pct_1 | pct_2 | p_val_adj |
|---|---|---|---|---|---|---|
| 0 | *MALAT1* | 6.57E-69 | -12.05 | 0.55 | 0.74 | 1.20E-64 |
| 0 | *CD74* | 1.63E-56 | -60.97 | 0.82 | 0.90 | 2.97E-52 |
| 0 | *EEF1A1* | 9.36E-55 | -21.51 | 0.74 | 0.84 | 1.70E-50 |
| 0 | *MMP3* | 3.91E-51 | -136.45 | 0.60 | 0.75 | 7.11E-47 |
| 0 | *ACTB* | 1.67E-49 | -19.56 | 0.80 | 0.88 | 3.05E-45 |
| 0 | *B2M* | 3.88E-48 | -70.56 | 0.94 | 0.96 | 7.05E-44 |
| 0 | *TMSB4X* | 3.90E-48 | -14.42 | 0.69 | 0.80 | 7.10E-44 |
| 0 | *COL3A1* | 6.07E-48 | -73.30 | 0.63 | 0.77 | 1.10E-43 |
| 0 | *FTH1* | 1.73E-47 | -23.46 | 0.67 | 0.79 | 3.15E-43 |
| 0 | *HLA-B* | 7.41E-47 | -70.56 | 0.88 | 0.93 | 1.35E-42 |
| 1 | *IGF1* | 1.24E-77 | 0.97 | 0.21 | 0.06 | 2.26E-73 |
| 1 | *COL1A1* | 8.88E-52 | 190.30 | 0.77 | 0.59 | 1.62E-47 |
| 1 | *MMP2* | 1.77E-30 | 25.89 | 0.56 | 0.40 | 3.22E-26 |
| 1 | *COL3A1* | 5.12E-24 | 61.33 | 0.82 | 0.72 | 9.31E-20 |
| 1 | *ELN* | 1.59E-22 | 7.58 | 0.23 | 0.13 | 2.90E-18 |
| 1 | *FNDC1* | 3.89E-19 | 0.31 | 0.11 | 0.05 | 7.08E-15 |
| 1 | *ASPN* | 4.95E-19 | 6.42 | 0.12 | 0.06 | 9.02E-15 |
| 1 | *AEBP1* | 8.85E-19 | 21.02 | 0.36 | 0.25 | 1.61E-14 |
| 1 | *COMP* | 3.25E-18 | 2.98 | 0.23 | 0.14 | 5.92E-14 |
| 1 | *CCL18* | 3.77E-16 | 5.23 | 0.33 | 0.23 | 6.86E-12 |
| 2 | *MMP3* | 2.67E-276 | 118.78 | 0.94 | 0.69 | 4.86E-272 |
| 2 | *MMP1* | 1.04E-210 | 36.51 | 0.72 | 0.28 | 1.89E-206 |
| 2 | *FN1* | 9.72E-146 | 269.70 | 0.99 | 0.98 | 1.77E-141 |
| 2 | *HTRA1* | 9.61E-138 | -22.09 | 0.88 | 0.72 | 1.75E-133 |
| 2 | *PRG4* | 7.54E-126 | Inf | 0.99 | 0.97 | 1.37E-121 |
| 2 | *CRTAC1* | 8.49E-125 | 18.19 | 0.85 | 0.67 | 1.54E-120 |
| 2 | *MT2A* | 3.02E-110 | 25.90 | 0.85 | 0.69 | 5.49E-106 |
| 2 | *CLU* | 5.26E-110 | -76.09 | 0.95 | 0.88 | 9.57E-106 |
| 2 | *VCAM1* | 9.61E-102 | 1.96 | 0.76 | 0.53 | 1.75E-97 |
| 2 | *PLA2G2A* | 5.94E-97 | 8.88 | 0.81 | 0.62 | 1.08E-92 |
| 3 | *MALAT1* | 1.21E-125 | -4.57 | 1.00 | 0.67 | 2.21E-121 |
| 3 | *ACTB* | 8.96E-66 | -34.59 | 0.77 | 0.88 | 1.63E-61 |
| 3 | *EEF1A1* | 6.41E-65 | -23.05 | 0.71 | 0.83 | 1.17E-60 |
| 3 | *HLA-B* | 1.12E-58 | -108.00 | 0.87 | 0.93 | 2.03E-54 |
| 3 | *RPL13A* | 4.02E-58 | -12.76 | 0.46 | 0.68 | 7.31E-54 |
| 3 | *TMSB4X* | 3.17E-57 | -27.81 | 0.65 | 0.79 | 5.77E-53 |
| 3 | *MT-CO3* | 5.39E-57 | -53.99 | 0.86 | 0.92 | 9.81E-53 |
| 3 | *TMSB10* | 1.31E-56 | -23.63 | 0.57 | 0.74 | 2.38E-52 |
| 3 | *MT-ND1* | 5.70E-56 | -20.54 | 0.71 | 0.84 | 1.04E-51 |
| 3 | *MT-ND4* | 2.13E-55 | -32.26 | 0.75 | 0.86 | 3.87E-51 |

| | | | | | | |
|---|---|---|---|---|---|---|
| 4 | *JCHAIN* | 7.99E-184 | 42.19 | 0.78 | 0.35 | 1.45E-179 |
| 4 | *IGH* | 2.73E-141 | Inf | 0.97 | 0.84 | 4.97E-137 |
| 4 | *MZB1* | 6.11E-97 | 2.10 | 0.49 | 0.19 | 1.11E-92 |
| 4 | *IGHGP* | 2.00E-92 | 74.42 | 0.42 | 0.15 | 3.63E-88 |
| 4 | *TXNDC5* | 5.11E-77 | 9.17 | 0.57 | 0.29 | 9.30E-73 |
| 4 | *DERL3* | 4.95E-72 | 0.67 | 0.26 | 0.07 | 9.02E-68 |
| 4 | *CD79A* | 4.75E-58 | 0.57 | 0.24 | 0.07 | 8.64E-54 |
| 4 | *XBP1* | 4.74E-57 | 4.20 | 0.47 | 0.24 | 8.62E-53 |
| 4 | *PIM2* | 1.03E-52 | 0.93 | 0.25 | 0.08 | 1.88E-48 |
| 4 | *SDC1* | 1.82E-50 | 0.93 | 0.23 | 0.07 | 3.31E-46 |
| 5 | *HLA-DQA1* | 5.13E-72 | 1.70 | 0.62 | 0.34 | 9.34E-68 |
| 5 | *LYZ* | 3.39E-66 | 4.87 | 0.68 | 0.42 | 6.17E-62 |
| 5 | *CD74* | 3.22E-56 | 2.16 | 0.94 | 0.88 | 5.86E-52 |
| 5 | *HLA-DRA* | 6.95E-44 | 31.21 | 0.90 | 0.87 | 1.26E-39 |
| 5 | *CXCL9* | 3.52E-41 | 2.41 | 0.41 | 0.21 | 6.41E-37 |
| 5 | *HLA-DRB1* | 2.76E-38 | 5.17 | 0.78 | 0.67 | 5.02E-34 |
| 5 | *FTH1* | 3.77E-37 | 1.21 | 0.84 | 0.76 | 6.86E-33 |
| 5 | *FTL* | 1.95E-36 | 2.99 | 0.90 | 0.86 | 3.55E-32 |
| 5 | *HLA-DPA1* | 2.04E-36 | 6.94 | 0.75 | 0.60 | 3.71E-32 |
| 5 | *TMSB10* | 4.28E-36 | 11.04 | 0.82 | 0.71 | 7.79E-32 |
| 6 | *IGFBP7* | 1.01E-164 | 2.96 | 0.82 | 0.40 | 1.84E-160 |
| 6 | *ACTA2* | 1.87E-148 | 4.21 | 0.48 | 0.12 | 3.40E-144 |
| 6 | *TAGLN* | 1.90E-112 | 1.93 | 0.42 | 0.12 | 3.46E-108 |
| 6 | *MCAM* | 7.17E-101 | 0.81 | 0.33 | 0.08 | 1.30E-96 |
| 6 | *VWF* | 1.60E-86 | 2.41 | 0.35 | 0.10 | 2.91E-82 |
| 6 | *MYL9* | 1.06E-73 | 2.30 | 0.64 | 0.34 | 1.92E-69 |
| 6 | *NR2F2* | 5.89E-58 | 0.61 | 0.20 | 0.05 | 1.07E-53 |
| 6 | *ACKR1* | 4.38E-49 | 0.64 | 0.19 | 0.05 | 7.97E-45 |
| 6 | *CCL19* | 4.31E-48 | 17.11 | 0.29 | 0.11 | 7.85E-44 |
| 6 | *NOTCH3* | 1.16E-40 | 0.71 | 0.30 | 0.12 | 2.12E-36 |
| 7 | *PRELP* | 9.81E-63 | 5.75 | 0.56 | 0.17 | 1.78E-58 |
| 7 | *HTRA1* | 2.30E-53 | 27.27 | 0.96 | 0.73 | 4.18E-49 |
| 7 | *PRG4* | 3.43E-26 | -Inf | 0.99 | 0.97 | 6.24E-22 |
| 7 | *CSN1S1* | 2.43E-22 | 5.88 | 0.25 | 0.08 | 4.43E-18 |
| 7 | *FN1* | 2.58E-22 | -264.68 | 0.99 | 0.98 | 4.70E-18 |
| 7 | *AMTN* | 2.65E-22 | 10.23 | 0.21 | 0.06 | 4.82E-18 |
| 7 | *CRTAC1* | 1.36E-21 | -13.01 | 0.80 | 0.69 | 2.48E-17 |
| 7 | *CLU* | 8.46E-20 | 81.27 | 0.90 | 0.89 | 1.54E-15 |
| 7 | *IGFBP5* | 1.34E-19 | -2.67 | 0.67 | 0.46 | 2.44E-15 |
| 7 | *SLC39A14* | 1.70E-19 | -0.47 | 0.47 | 0.24 | 3.10E-15 |
| 8 | *FABP4* | 1.51E-47 | 5.49 | 0.26 | 0.05 | 2.75E-43 |

| | | | | | | |
|---|---|---|---|---|---|---|
| 8 | *PLIN4* | 1.03E-41 | 0.42 | 0.19 | 0.03 | 1.88E-37 |
| 8 | *LPL* | 2.04E-37 | 0.79 | 0.15 | 0.02 | 3.71E-33 |
| 8 | *C3* | 1.15E-33 | -0.51 | 0.59 | 0.27 | 2.09E-29 |
| 8 | *SCD* | 1.48E-23 | 0.74 | 0.26 | 0.08 | 2.70E-19 |
| 8 | *SAA1* | 6.65E-19 | 0.86 | 0.18 | 0.05 | 1.21E-14 |
| 8 | *B2M* | 4.73E-12 | -155.66 | 0.96 | 0.96 | 8.60E-08 |
| 8 | *HLA-A* | 8.23E-11 | -65.66 | 0.69 | 0.80 | 1.50E-06 |
| 8 | *HLA-B* | 1.13E-10 | -119.66 | 0.91 | 0.92 | 2.05E-06 |
| 8 | *HLA-E* | 6.72E-10 | -4.90 | 0.56 | 0.68 | 1.22E-05 |
| 9 | *HBA2* | 0.00E+00 | 2.00 | 1.00 | 0.05 | 0.00E+00 |
| 9 | *HBA1* | 7.79E-33 | 1.84 | 0.14 | 0.02 | 1.42E-28 |
| 9 | *MMP2* | 1.82E-08 | -26.12 | 0.24 | 0.43 | 3.32E-04 |
| 9 | *CLU* | 3.92E-08 | -149.47 | 0.78 | 0.89 | 7.14E-04 |
| 9 | *CD68* | 8.59E-08 | -4.06 | 0.19 | 0.36 | 1.56E-03 |
| 9 | *ENO1* | 1.02E-07 | -4.21 | 0.26 | 0.44 | 1.86E-03 |
| 9 | *PSAP* | 1.39E-07 | -24.50 | 0.72 | 0.81 | 2.53E-03 |
| 9 | *SAT1* | 1.73E-07 | -4.67 | 0.29 | 0.46 | 3.16E-03 |
| 9 | *CTSB* | 2.22E-07 | -19.17 | 0.67 | 0.77 | 4.04E-03 |
| 9 | *HTRA1* | 2.70E-07 | -96.38 | 0.66 | 0.74 | 4.91E-03 |

Table S2. Top 10 differentially expressed genes for 10 spatial transcriptomics clusters are shown.

| | OR | p-value |
|---|---|---|
| Naïve B cells (SC-B1) | 20.3 | <2.2e-16 |
| Memory B cells (SC-B2) | 99.6 | <2.2e-16 |
| Plasma cells (SC-B4) | 3.27 | <2.2e-16 |

Table S3. Fisher's exact test for memory B cells and the naïve B cells, memory B cells and plasma cells clusters from previously published studies *(8)* is shown.

| | OR | p-value |
|---|---|---|
| Naïve B cells (SC-B1) | 0.438 | 1 |
| Memory B cells (SC-B2) | 0 | 1 |
| Plasma cells (SC-B4) | 45.1 | <2.2e-16 |

Table S4. Fisher's exact test for plasma cells and the naïve B cells, memory B cells and plasma cell clusters from previously published studies *(8)* is shown.
